# Is there a causal relationship between executive function and liability to mental health and substance use? A Mendelian randomisation approach

**DOI:** 10.1101/2022.01.12.22269153

**Authors:** Sabrina M. I. Burton, Hannah M. Sallis, Alexander S. Hatoum, Marcus R. Munafò, Zoe E. Reed

**Affiliations:** School of Psychological Science, University of Bristol, Bristol, UK; MRC Integrative Epidemiology Unit at the University of Bristol, Bristol, UK; Centre for Academic Mental Health, Population Health Sciences, University of Bristol, Bristol, UK; Department of Psychiatry, Washington University School of Medicine, St Louis, MO, USA; National Institute for Health Research Bristol Biomedical Research Centre, University Hospitals Bristol NHS Foundation Trust and University of Bristol, Bristol, UK

## Abstract

**Background:** Executive function consists of several cognitive control processes that are able to regulate lower level processes. Poorer performance in tasks designed to test executive function is associated with a range of psychopathologies such as schizophrenia, major depressive disorder (MDD) and anxiety, as well as with smoking and alcohol consumption. Despite these well-documented associations, whether they reflect causal relationships, and if so in what direction, remains unclear. We aimed to establish whether there is a causal relationship between a latent factor for performance on multiple executive function tasks – which we refer to as common executive function (cEF) – and liability to schizophrenia, MDD, anxiety, smoking initiation, alcohol consumption, alcohol dependence and cannabis use disorder (CUD), and the directionality of any relationship observed.

**Methods:** We used a two-sample bidirectional Mendelian randomisation (MR) approach using genome-wide association study (GWAS) summary data from large cohorts (N=17,310 to 848,460) to examine whether causal relationships exist, and if so in which direction.

**Results:** We found evidence of a causal effect of increased cEF on reduced schizophrenia liability (IVW: OR=0.10; 95% CI 0.05 to 0.19; p-value=3.43×10^−12^), reduced MDD liability (IVW: OR=0.52; 95% CI 0.38 to 0.72; p-value=5.23×10^−05^), decreased drinks per week (IVW: β=−0.06; 95% CI −0.10 to −0.02; p-value=0.003), and reduced CUD liability (IVW: OR=0.27; 95% CI 0.12 to 0.61; p-value=1.58×10^−03^). We also found evidence of a causal effect of increased schizophrenia liability on decreased cEF (IVW: β=−0.04; 95% CI −0.04 to −0.03; p-value=3.25×10^−27^), as well as smoking initiation on decreased cEF (IVW: β=−0.06; 95%CI −0.09 to −0.03; p-value=6.11×10^−05^).

**Conclusion:** Our results indicate a potential bidirectional causal relationship between a latent factor measure of executive function (cEF) and schizophrenia liability, a possible causal effect of increased cEF on reduced MDD liability, CUD liability, and alcohol consumption, and a possible causal effect of smoking initiation on decreased cEF. These results suggest that executive function should be considered as a potential risk factor for some mental health and substance use outcomes, and may also be impacted by mental health (particularly schizophrenia). Further studies are required to improve our understanding of the underlying mechanisms of these effects, but our results suggest that executive function may be a promising intervention target. These results may therefore inform the prioritisation of experimental medicine studies (e.g., of executive function interventions), for both mental health and substance use outcomes, to improve the likelihood of successful translation.

## Introduction

The ability to perform nearly all of the activities required for daily living is mediated by executive function (EF)^1^ – the ability to perform self-directed behaviour toward a goal and to enable self-regulation. The prefrontal cortex is the neural substrate of EF, including cognitive control functions that regulate lower-level processes such as decision making^1^. There are different aspects of EF, including inhibitory control, working memory and task switching.

EF is thought to be impaired in individuals with a range of mental health problems^1^, and there is evidence that it is also associated with substance use^2^. For example, poorer EF has been observed among individuals with schizophrenia^1,3–5^, major depressive disorder (MDD)^6,7^, and anxiety^8,9^, as well as in people who smoke both cigarettes^10,11^ and cannabis^12,13^ and consume alcohol^10,14^. The direction of association between EF and these phenotypes is unclear^1^, with some studies suggesting EF deficits prior to these^2^ and others suggesting they occur after^3^. It is unclear whether these associations represent causal pathways, and if so what the direction of any causal effect might be. EF is potentially modifiable^15^, and drug repurposing analyses for a common EF factor (cEF) score^16^ have suggested that this cEF may also be modifiable. Therefore, if we can better understand the relationship between EF and mental health and substance use outcomes, this will help to inform intervention development.

Mendelian randomisation (MR) is a well-established method for causal inference, which relies on approximations of Mendel’s laws of segregation and random assortment^17^. MR is based on instrumental variable (IV) analysis, with single nucleotide polymorphisms (SNPs) that are robustly associated with the exposure used as IVs. MR is subject to three core assumptions: i) the genetic instrument is robustly associated with the exposure of interest (relevance), ii) there is no confounding of the genetic instrument and the outcome (independence), and iii) the genetic instrument only influences the outcome via the exposure (exclusion restriction). There are different MR methods that test potential violations of these assumptions and therefore a consistent effect estimate across different approaches would provide greater evidence of a truly causal effect, robust to the assumptions of MR. MR minimises the effect of confounding variables as the genetic variants are randomly assigned at conception^18^. It also overcomes issues around reverse causation as these genetic variants precede any outcomes^19^.

Summary data from genome-wide association studies (GWAS) can be used as the genetic instruments in MR. A recent GWAS of cEF score, conducted in the UK Biobank (European ancestry), identified 90 genome-wide significant hits^16^. The factor score was created from five different EF tasks – trail making, symbol-digit substitution, digit span, prospective memory, and pairs memory – using confirmatory factor analysis. Unlike, previous studies focusing on specific EF tasks, the cEF incorporates multiple facets which may better capture the cognitive component of psychopathology. In particular, single executive function tasks are noisy measures of executive functioning, with large method variance components reflecting lower level cognitive processes (the “task impurity problem” ^20^). By combining multiple tasks, we can create a more “pure” measure of executive functioning. Further, past work has shown that this common component is correlated with – but separable from – IQ, and is genetically associated with psychopathology over and above the genetic influence of other cognitive factors^16^.

We examined whether there were causal relationships between EF and a range of mental health and substance use phenotypes, and the direction of any effect – for example, does poor mental health lead to poorer EF, or vice versa? We did this by applying a two-sample MR approach, where the SNP-exposure and SNP-outcome estimates are obtained from genome-wide association studies (GWAS) in independent samples and used to estimate causal effects. We focused on schizophrenia, MDD, anxiety, smoking initiation, alcohol consumption (drinks per week), alcohol dependence, and cannabis use disorder (CUD), using a bidirectional approach to determine the causal direction of these relationships.

## Methods

### Data Sources

We used GWAS data from several studies, shown in Table 1. To minimise sample overlap, we excluded some samples that contributed to the original GWAS in our analyses, as indicated in Table 1.

**Table 1.** GWAS for executive function, mental health and substance use outcomes.

| Phenotype | Author | Sample | Final N |
| --- | --- | --- | --- |
| Executive function | Hatoum et al, 2020<sup>16</sup> | UKBB | 427,037 |
| Schizophrenia | Schizophrenia Working Group of the PGC, 2020<sup>21</sup> | PGC | Cases = 69,369<br>Controls = 236,642 |
| Major depressive disorder | Wray et al, 2018<sup>22</sup> | PGC, excluding UKBB and 23andMe | Cases = 45,396<br>Controls = 97,250 |
| Anxiety | Otowa et al, 2016<sup>23</sup> | ANGST | Cases = 5,712<br>Controls = 11,598 |
| Smoking initiation | Liu et al, 2019<sup>24</sup> | GSCAN, excluding UKBB | 848,460 |
| Drinks per week | Liu et al, 2019<sup>24</sup> | GSCAN, excluding UKBB | 630,154 |
| Alcohol dependence | Walters et al 2018<sup>25</sup> | PGC Substance Use Disorders working group | Cases = 8,485<br>Controls = 20,272 |
| Cannabis use disorder | Johnson et al, 2020<sup>26</sup> | PGC Substance Use Disorders working group, iPSYCH and deCODE | Cases = 14,080<br>Controls = 343,736 |

UKBB= UK Biobank, PGC= Psychiatric Genomics Consortium, ANGST= Anxiety NeuroGenetics Study,
GSCAN= GWAS and Sequencing Consortium of Alcohol and Nicotine use

#### Executive function

We used summary data from the most recent GWAS of cEF^16^, which identified 90 independent genome-wide significant (p<5×10^−08^) SNPs associated with a cEF score, where a higher score reflects increased EF.

#### Schizophrenia

We used summary data from the most recent Psychiatric Genomics Consortium (PGC) GWAS of schizophrenia^21^, which identified 294 independent genome-wide significant SNPs. Cases mostly included participants diagnosed with schizophrenia (although diagnoses of other psychotic disorders were also included in some samples).

#### Major depressive disorder (MDD)

We used summary data from the most recent PGC GWAS of MDD^22^, which identified 44 independent genome-wide significant SNPs. Cases of MDD were either diagnosed by a clinical professional, or through structured interviews with trained interviewers, using the DSM IV, ICD-9 or ICD-10 criteria.

#### Anxiety

We used summary data from a GWAS of anxiety^23^, which identified one independent genome-wide significant SNP. In this meta-analysis, there were up to five different anxiety disorder phenotypes included in the nine samples from seven independent cohorts. Lifetime DSM-based anxiety disorder diagnostic assessments were available for all cohorts except for the Rotterdam study, in which only one-year prevalence was assessed. Each study assessed DSM-based criteria for the following six-lifetime clinical phenotypes: generalised anxiety disorder (GAD), panic disorder, agoraphobia, social phobia, specific phobia and MDD, however, any subject reporting a mood disorder only was removed from analyses.

#### Smoking initiation

We used summary data from the most recent GWAS of smoking initiation^24^, which identified 378 conditionally independent genome-wide significant SNPs associated with ever being a regular smoker (current or former). Participants were asked whether they had smoked more than 100 cigarettes in their lifetime and whether they had ever smoked every day for at least a month or ever smoked regularly. To obtain summary statistics for the full sample included in the GWAS and Sequencing Consortium of Alcohol and Nicotine use (GSCAN) GWAS excluding UK Biobank data, we meta-analysed results from GWAS of 23andMe, Inc. only data and all results excluding UK Biobank and 23andMe. The meta-analysis was conducted using the genome-wide association meta-analysis (GWAMA) software^27^.

#### Drinks per week

We used summary data from the most recent GWAS of drinks per week^24^, which identified 99 conditionally independent genome-wide significant SNPs associated with the average number of drinks a participant reported drinking each week. Participants were asked about the number of alcoholic beverages they had in the past week and the average number of drinks per week than they had in the past year. Data were log-transformed prior to the GWAS. This measure did not account for the type of alcohol consumed and for any study with ranges, the mid-range value was used. Again, to obtain summary statistics for the full GSCAN GWAS excluding UK Biobank data, we meta-analysed results from GWAS of 23andMe only data and all results excluding UK Biobank and 23andMe.

#### Alcohol Dependence

We used summary data from the PGC substance use disorders working group GWAS of alcohol dependence^25^, which identified one conditionally independent genome-wide significant SNPs in their European GWAS associated with alcohol dependence. Alcohol dependence cases were those that met criteria for DSM-IV or DSM-III-R alcohol dependence diagnosis.

#### Cannabis Use Disorder (CUD)

We used summary data from the most recent GWAS of CUD^26^ from the PGC substance use disorders working group, iPSYCH and deCODE which identified 2 conditionally independent genome-wide significant SNPs associated with CUD. CUD cases were those that met criteria for DSM-5, DSM-IV, DSM-III-R or ICD-10 cannabis abuse or dependence.

### Statistical analyses

The analysis plan for this study was pre-registered on the Open Science Framework (https://osf.io/j3tb5). We conducted two-sample MR analyses using R (version 4.0.3)^28^ and the package TwoSampleMR (version 0.5.0)^29,30^. In order to assess potential bidirectional pathways, we conducted two-sample MR analyses with cEF as the exposure for one direction, and as the outcome in the other direction (see Figure 1), when assessing causal relationships with liability to schizophrenia, MDD, anxiety, smoking initiation, drinks per week, alcohol dependence and CUD.

**Figure 1.**
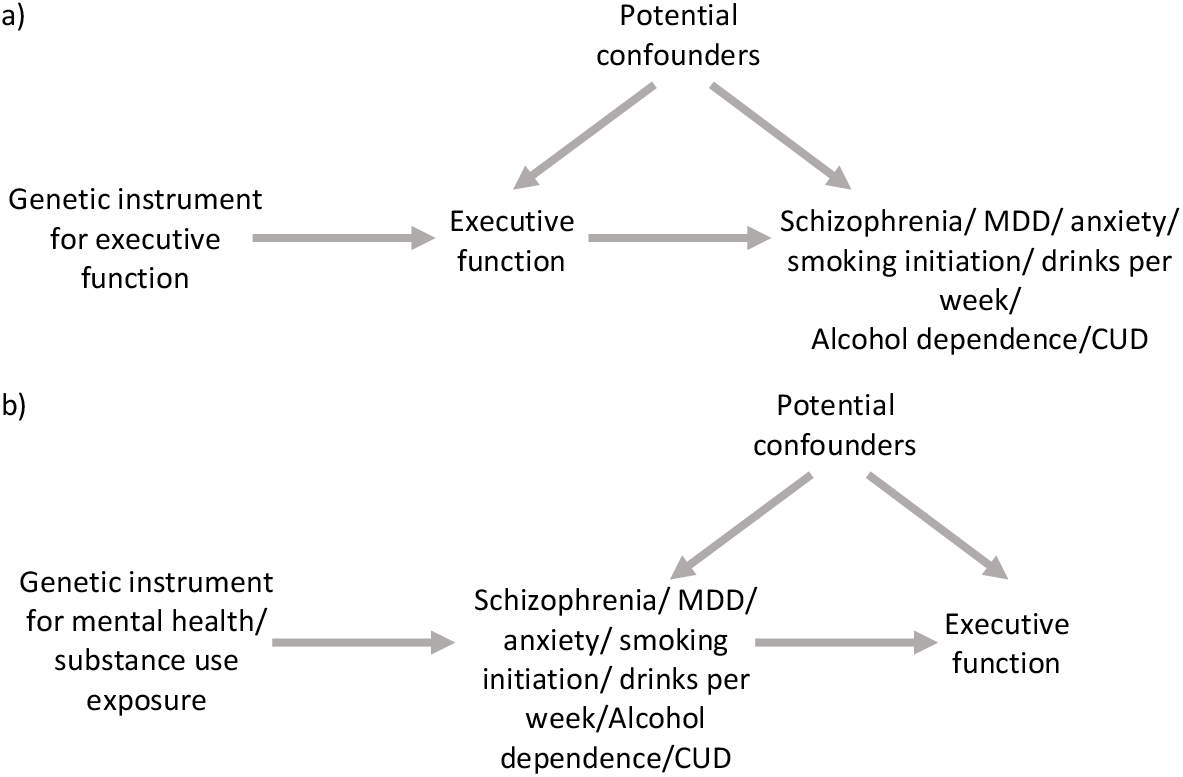
Bidirectional two-sample Mendelian randomisation between a common executive function factor score and liability to mental health and substance use outcomes. Directed acyclic graph for the potential causal effects being examined with a) executive function as the exposure and liability to mental health and substance use phenotypes as the outcomes and b) liability to mental health and substance use phenotypes as the exposures and executive function as the outcome. MDD=Major depressive disorder, CUD=Cannabis use disorder

We used independent genome-wide significant SNPs for the exposure of interest as instruments in the MR analyses, except when anxiety, alcohol dependence and CUD were the exposures, where we used a p-value threshold of 1×10^−05^ to select SNPs due to the low number of genome-wide significant SNPs. We excluded all SNPs in linkage disequilibrium (LD) using an r^2^ of 0.001, a window of 10000 kb and the European 1000 genomes reference panel. Where there were palindromic SNPs, we tried to infer the positive strand based on allele frequencies, but if this was not possible, then these SNPs were also excluded. Where an exposure SNP was not available in the outcome data, we attempted to identify a suitable proxy SNP using the LDproxy tool from LDlink^31^. We pruned the SNPs extracted to r^2^≥0.8 and extracted the first SNP that was present in the exposure and outcome data. After exclusions and identifying any proxy SNPs we searched for the remaining cEF SNPs in the outcome GWAS (73 for schizophrenia, 85 for MDD, 83 for anxiety, 82 for smoking initiation, 83 for drinks per week, 73 for alcohol dependence and 73 for CUD) and the remaining mental health and substance use exposure SNPs in the cEF outcome (175 for schizophrenia, 29 for MDD, 17 for anxiety, 187 for smoking initiation, 67 for drinks per week, 19 for alcohol dependence and 37 for CUD). The GWAS summary statistics for the exposure and outcome in each analysis were harmonised so that the SNP allele-exposure and SNP allele-outcome associations were in the same direction.

We used several different MR methods to assess these putative causal relationships: inverse-variance weighted (IVW)^32^, MR-Egger^33^, weighted median^34^, simple mode and weighted mode^35^, and Steiger filtering^36^ MR methods. We used the IVW approach as our main method with the other methods used as sensitivity analyses.

The IVW approach constrains the intercept to pass through zero, assuming no horizontal pleiotropy. We tested for heterogeneity between the individual SNPs included in the genetic instrument using Cochran’s test of heterogeneity. The MR-Egger method tests for overall directional pleiotropy by not constraining the intercept to pass through zero. If the intercept is not zero then this is indicative of directional horizontal pleiotropy. We also assessed heterogeneity between the individual SNPs whilst adjusting for any directional pleiotropy for the MR-Egger method using Rucker’s Q test. We used the weighted median method to obtain estimates under the assumption that at least 50% of the SNPs satisfy the MR assumptions and are valid IVs. Finally, we used the mode-based approaches to obtain estimates for the largest cluster of SNPs, where SNPs not in that cluster could be invalid. The weighted method accounts for the largest weights of SNPs. We also conducted single SNP and leave-one-out analyses.

Where we found evidence of a bidirectional causal relationship, we ran Steiger filtering. This allows orientation of the direction of effect where the underlying biology of genetic variants is less clear, by identifying which SNPs explain more variance in the outcome than the exposure and then repeating the MR analyses excluding those SNPs to rule out reverse causation^36^.

In cases where we found evidence for a causal effect between a given exposure and outcome, we present plots of these results in the Supplementary Material. These plots include scatter plots of the SNP-exposure and SNP-outcome associations with the causal effect estimates from each MR method presented, forest plots for causal effects of each SNP in the instrument (which can indicate if heterogeneity is present), plots presenting the leave-one-out results and funnel plots of each SNP included in the instrument, where a symmetrical plot indicates that the effects of each SNP included are similar to the average effect and asymmetry may indicate horizontal pleiotropy.

We calculated weighted and unweighted regression dilution I-squared statistics for each analysis^37^, presented in Supplementary Table S1, which give an indication of the amount of bias in the ‘NO Measurement Error’ (NOME) assumption in the MR-Egger estimate^34^. If the I-squared statistic is 0.9 or above this indicates minimal bias in the MR-Egger estimate and therefore we present the MR-Egger results for these associations. If either the weighted or unweighted I-squared statistics were between 0.6 and 0.9, this may indicate regression dilution bias and therefore we ran simulation extrapolation (SIMEX) corrections, to obtain bias-adjusted point estimates for MR-Egger and we present these results in place of MR-Egger. Anything below 0.6 means that the bias may be too large and therefore we do not report either the SIMEX correction or the MR-Egger results. We also estimated the mean F-statistic for each analysis, indicative of instrument strength, where a value under 10 may indicate a weak instrument^37^.

## Data availability

The data used in this study are publicly-available GWAS data for cEF (available upon request by contacting the authors), schizophrenia (https://www.med.unc.edu/pgc/download-results/scz/), MDD (https://www.med.unc.edu/pgc/download-results/mdd/), anxiety (https://www.med.unc.edu/pgc/download-results/angst/?choice=Other+GWAS+DataAnxiety+Neuro+Genetics+Study+%28ANGST%29), smoking initiation and drinks per week (data with UK Biobank and 23andMe removed can be found here: https://conservancy.umn.edu/handle/11299/201564, and for 23andMe data access needs to be requested (see below), alcohol dependence (https://figshare.com/articles/dataset/sud2018-alc/14672187) and CUD (https://figshare.com/articles/dataset/sud2020-cud/14842692). The full GWAS summary statistics for the 23andMe discovery data set will be made available through 23andMe to qualified researchers under an agreement with 23andMe that protects the privacy of the 23andMe participants. Please visit https://research.23andme.com/collaborate/#dataset-access/ for more information and to apply to access the data.

## Code availability

The analysis code that forms the basis of the results presented here is available from the University of Bristol’s Research Data Repository (http://data.bris.ac.uk/data/), DOI : To be made available upon publication).

## Results

Our two-sample MR results for the causal effects of cEF on mental health and substance use outcomes are presented in Table 2.

**Table 2.**
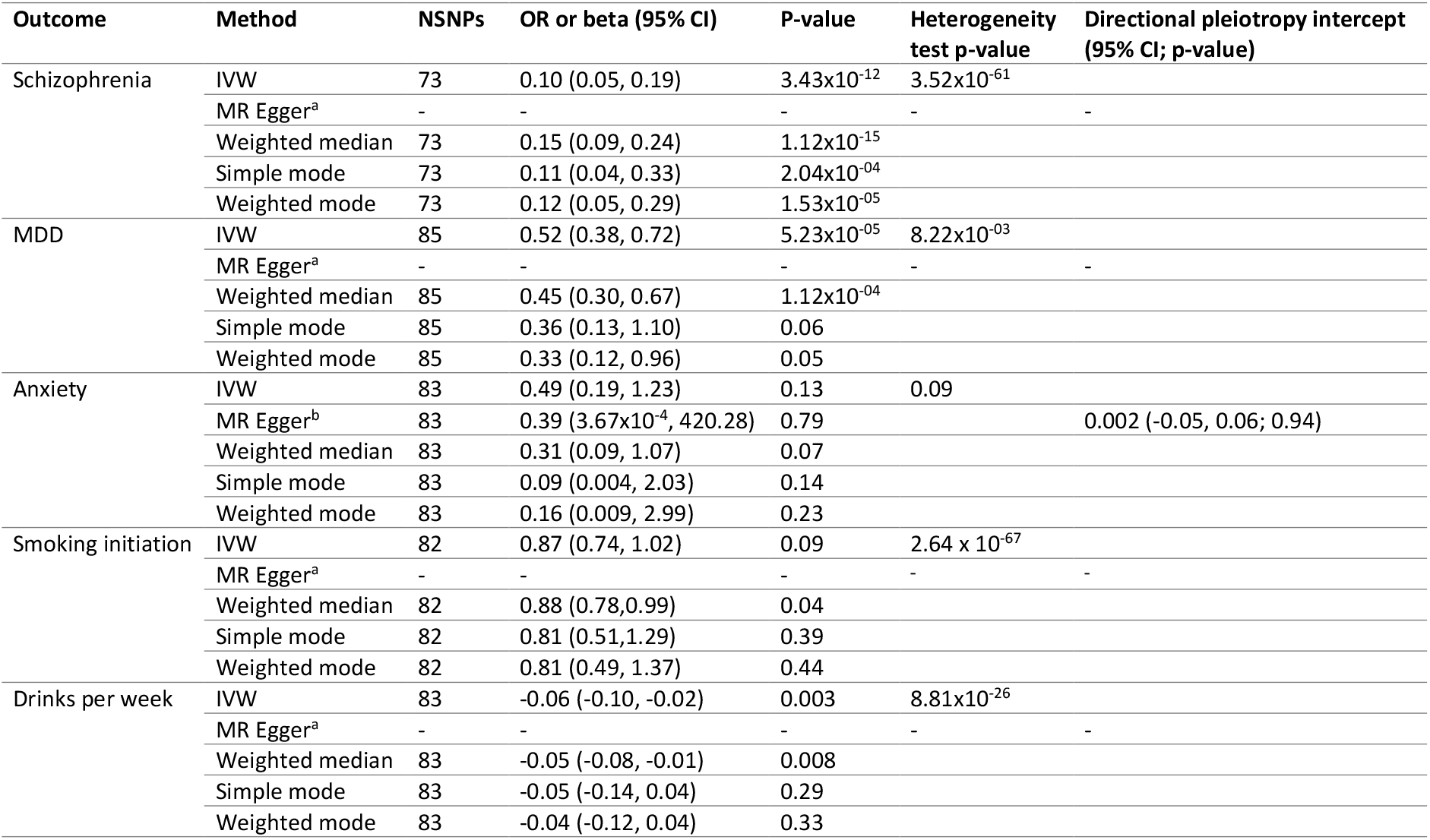
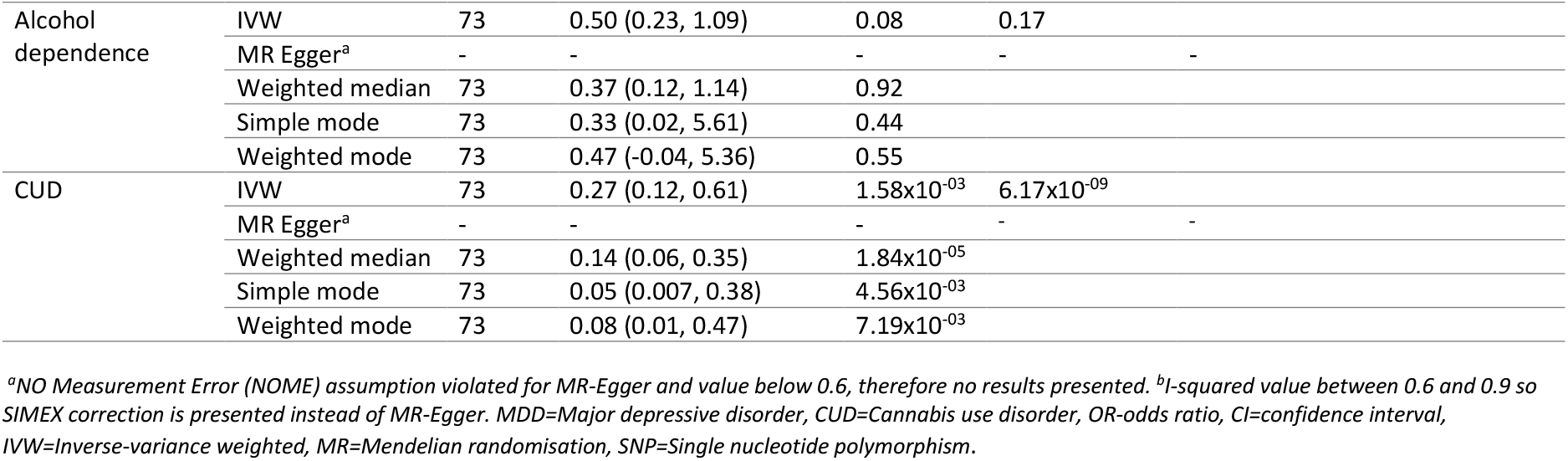
Two-sample Mendelian randomisation results with executive function (cEF) as the exposure.

### Schizophrenia

We found strong evidence of a causal effect of increased cEF on reduced odds of schizophrenia (IVW: OR=0.10; 95% CI 0.05 to 0.19; p-value=3.43×10^−12^) for all methods except MR Egger, which we were unable to estimate due to violation of the NOME assumption (See Supplementary Table S1). These results were in a consistent direction across the different MR analyses (Supplementary Figure S1). However, we did observe evidence of heterogeneity for the IVW estimate (Supplementary Figure S2) and some asymmetry in the funnel plot (Supplementary Figure S3), although our leave one out analyses did not indicate that a single SNP was driving the association (Supplementary Figure S4). Steiger filtering indicated that only 41% of SNPs instrumenting cEF explained more variance in cEF than schizophrenia, and results were attenuated when repeating analyses with this subset of SNPs. However, these results were in the same direction as the main results (Supplementary Table 2).

### Major depressive disorder

We found strong evidence of a causal effect of increased cEF on reduced odds of MDD (IVW: OR=0.52; 95% CI 0.38 to 0.72; p-value=5.23×10^−05^). These results were in a consistent direction across the different MR analyses (Supplementary Figure S5), and there was evidence of a causal effect for all methods except simple mode and MR Egger, which again we unable to estimate due to violation of the NOME assumption. Here we also found evidence of heterogeneity for the IVW estimate (Supplementary Figure S6), and some asymmetry in the funnel plot (Supplementary Figure S7); however, leave one out analyses did not indicate that a single SNP was driving the association (Supplementary Figure S8). Steiger filtering indicated that 81% of SNPs instrumenting cEF explained more variance in cEF than MDD, but again results were similar to the main results when using this subset of SNPs (Supplementary Table 2).

### Anxiety

We did not find evidence of a causal effect of cEF on anxiety liability (IVW: OR=0.49; 95% CI 0.19 to 1.23; p-value=0.13).

### Smoking initiation

We did not find clear evidence of a causal effect of cEF on smoking initiation (IVW: OR=0.87; 95% CI 0.74 to 1.02; p-value=0.09) for any of the MR analyses.

### Drinks per week

We found some evidence of a causal effect of increased cEF on decreased number of alcoholic drinks per week consumed (IVW: β=−0.06; 95% CI −0.10 to −0.02; p-value=0.003). These results were in a consistent direction across the different MR analyses (Supplementary Figure S9), although there was only evidence of a causal effect for the IVW method. We also found evidence of heterogeneity here (Supplementary Figure S10) and slight asymmetry in the funnel plot (Supplementary Figure S11); however, leave one out analyses did not indicate that a single SNP was driving the association (Supplementary Figure S12).

### Alcohol dependence

We did not find clear evidence of a causal effect of cEF on alcohol dependence liability (IVW: OR=0.50; 95% CI 0.23 to 1.28; p-value=0.08) for any of the MR analyses.

### Cannabis Use Disorder (CUD)

We found strong evidence of a causal effect of increased cEF on reduced odds of CUD (IVW: OR=0.27; 95% CI 0.12 to 0.61; p-value=1.58×10^−03^) for all methods except MR Egger, which we were unable to estimate due to violation of the NOME assumption (See Supplementary Table S1). These results were in a consistent direction across the different MR analyses (Supplementary Figure S13). However, we did observe evidence of heterogeneity for the IVW estimate (Supplementary Figure S14) and slight asymmetry in the funnel plot (Supplementary Figure S15), although our leave one out analyses did not indicate that a single SNP was driving the association (Supplementary Figure S16).

Our two-sample MR results for causal effects of the mental health and substance use phenotypes on cEF are presented in Table 3.

**Table 3.**
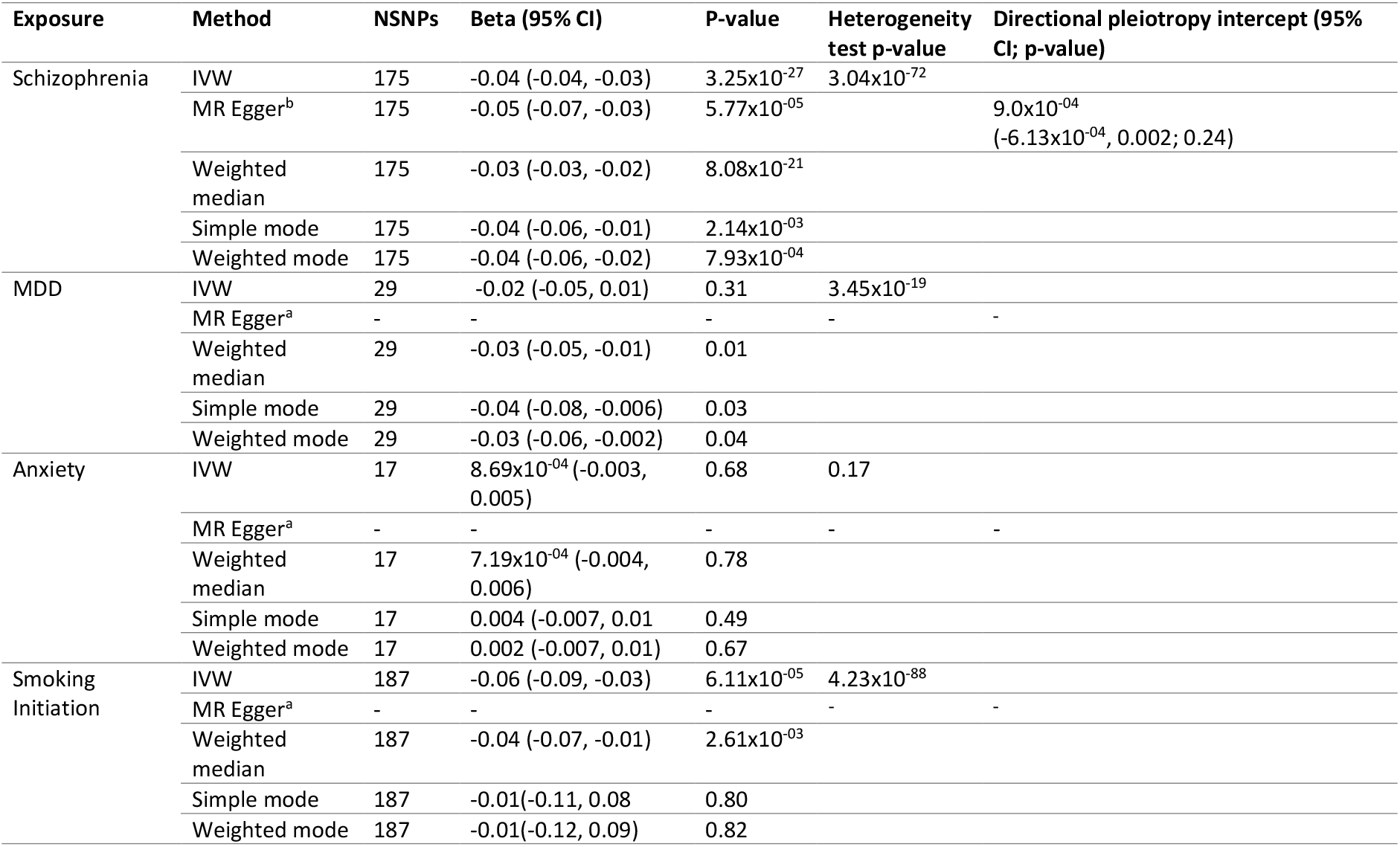
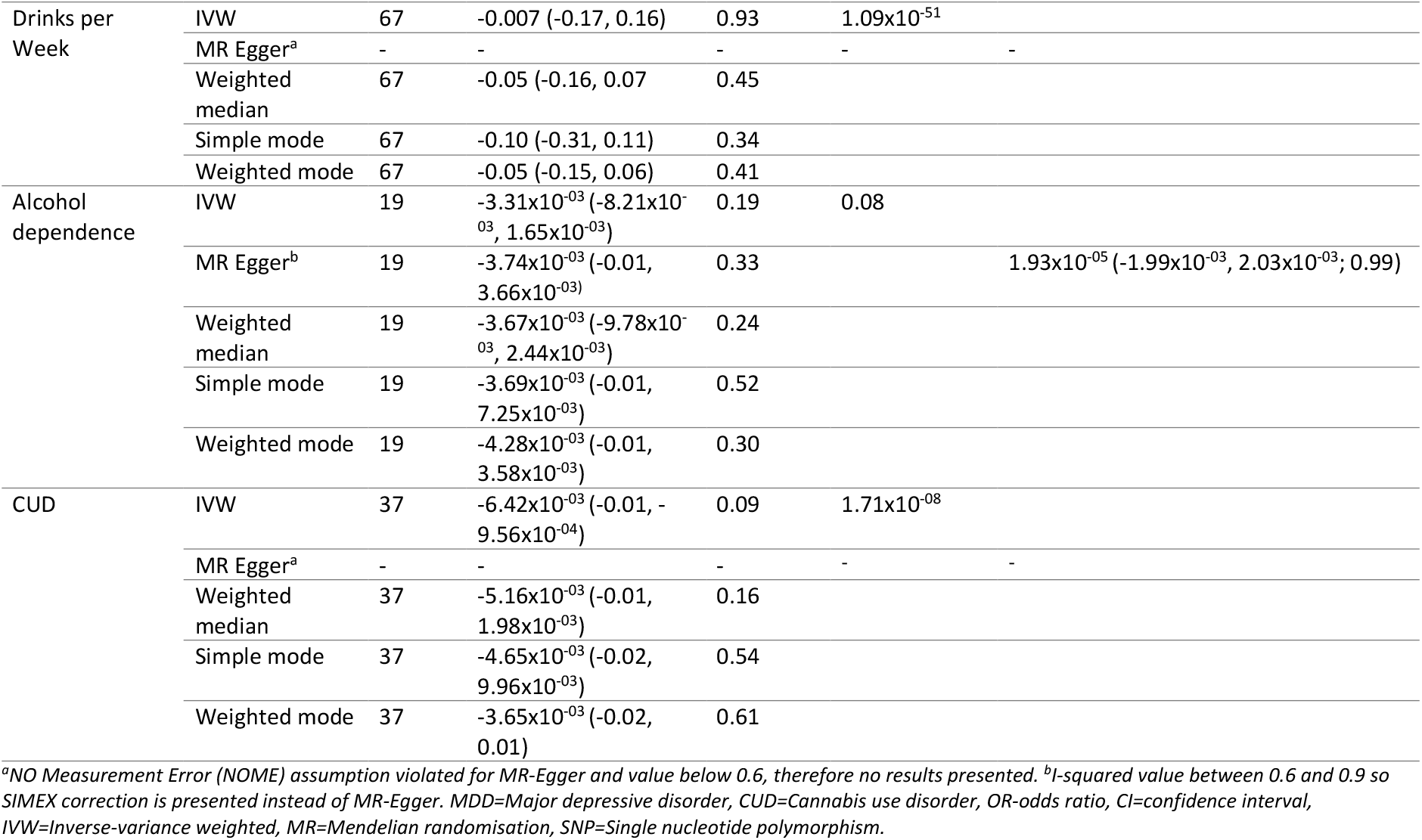
Two-sample Mendelian randomisation results with executive function (cEF) as the outcome.

### Schizophrenia

We found strong evidence of a causal effect of increased odds of schizophrenia on decreased cEF (IVW: β=−0.04; 95% CI −0.04 to −0.03; p-value=3.25×10^−27^). These results were in a consistent direction across the different MR analyses (Supplementary Figure S17). However, we did observe evidence of heterogeneity for the IVW estimate (Supplementary Figure S18) and some asymmetry in the funnel plot (Supplementary Figure S19), although there was little evidence of directional pleiotropy from the SIMEX estimate and our leave one out analyses did not indicate that a single SNP was driving the association (Supplementary Figure S20). Steiger filtering indicated that all SNPs instrumenting schizophrenia explained more variance in schizophrenia than cEF; therefore these analyses were not repeated (Supplementary Table 2).

### MDD

We did not find evidence of a causal effect of MDD liability on cEF (IVW: β=−0.02; 95% CI −0.05 to 0.01; p-value=0.31) using IVW. However, evidence was stronger using other sensitivity methods and the direction of effect was consistent across all approaches i.e., for increased odds of MDD on decreased cEF. Steiger filtering indicated that all SNPs instrumenting schizophrenia explained more variance in MDD than cEF; therefore these analyses were not repeated (Supplementary Table 2).

### Anxiety

We did not find evidence of a causal effect of anxiety liability on cEF (IVW: β=8.69×10^−04^; 95% CI −0.003 to 0.005; p-value=0.68) for any of the MR analyses.

### Smoking initiation

We found strong evidence of a causal effect of smoking initiation on decreased cEF (IVW: β=−0.06; 95% CI −0.09 to −0.03; p-value=6.11×10^−05^) and there was evidence of this causal effect using the weighted median method but not the other MR methods, although the direction of effect was consistent (Supplementary Figure S21). We did find evidence of heterogeneity for the IVW estimate (Supplementary Figure S22) and slight asymmetry in the funnel plot (Supplementary Figure S23); however, our leave one out analyses did not indicate that a single SNP was driving the association (Supplementary Figure S24).

### Drinks per week

We did not find clear evidence of a causal effect of cEF on number of alcoholic drinks per week consumed (IVW: β=−0.007; 95% CI −0.17 to 0.16; p-value=0.93).

### Alcohol dependence

We did not find clear evidence of a causal effect of alcohol dependence liability on cEF (IVW: β=−3.31×10^−03^; 95% CI −8.21×10^−03^ to 1.65×10^−03^; p-value=0.09) for any of the MR analyses.

### Cannabis Use Disorder (CUD)

We did not find clear evidence of a causal effect of CUD liability on cEF (IVW: β=−6.42×10^−03^; 95% CI −0.01 to 9.56×10^−04^; p-value=0.09) for any of the MR analyses.

## Discussion

We examined whether there was evidence of causal effects of cEF on schizophrenia, MDD, anxiety, smoking initiation, drinks per week, alcohol dependence and CUD. We also examined the reverse direction (i.e., causal effects of mental health and substance use on cEF). Evidence of a causal effect in both directions may be indicative of a bidirectional relationship or some other underlying common risk factor.

Our main findings were evidence of a causal relationship between increased cEF and reduced schizophrenia liability in both directions. Steiger filtering supported the finding of bidirectional effects, despite some attenuation of results for cEF on schizophrenia. This causal effect supports previous observational studies, which have found that people with schizophrenia have poorer EF^1,3–5^; however, our MR analyses provide evidence that these associations may reflect causal pathways. The fact that we find causal effects between schizophrenia liability and cEF in both directions may point to this association being bidirectional or due to an underlying common risk factor.

The observed causal effect of increased cEF on reduced MDD liability is also interesting, as previous studies have provided mixed evidence for the directionality of this relationship^6,7^. We did not find strong evidence of a causal effect of MDD on cEF using the IVW approach; however, evidence was stronger when using other MR methods, and the direction of effect we observed was negative and consistent across these. It may also be possible that there is lower power in the MDD instrument than the cEF instrument, due to the lower number of SNPs, which may also be why we do not observe a consistent effect of MDD on cEF. Therefore, we cannot rule out the possibility of a bidirectional relationship and Steiger filtering suggested that the effects could be bidirectional as well. We cannot draw any strong conclusions from our results regarding anxiety and cEF. Whilst we do not find evidence of a causal relationship, this does not mean that there is definitely no effect, but rather that our study may have lacked the power to detect a causal effect here, particularly given that we find some evidence of a possible causal effect of MDD on cEF in our sensitivity analyses and previous studies have reported high genetic correlations between MDD and anxiety^38^. Thus the association found in previous studies needs further investigation still^8,9^.

Finally, we also found some evidence of a causal effect of smoking initiation on decreased cEF and increased cEF on decreased drinks per week and reduced CUD liability, all in a consistent direction with previous observational studies^10–14^. However, evidence for the smoking and drinks per week findings was weak, so further studies examining this would be useful.

Our results are in contrast to a previous study which used latent causal variable (LCV) analyses and did not find evidence of any causal effects of cEF on schizophrenia, MDD, anxiety, alcohol use disorder or other traits examined^16^. LCV analysis relies on the assumption that there is a latent variable that mediates the genetic correlation between two traits and uses whole genome summary statistics to estimate genetic causality. One trait is partially genetically causal for the other if it is strongly genetically correlated with the LCV. However, LCV aims to capture the overall direction of causality and therefore may be less appropriate for relationships where a bidirectional effect may be present, as we observe in our study. Whereas MR does allow for for bidirectional relationships^39^. Although, LCV has better control for pleiotropy^16^, meaning the difference between our results and those in the previous LCV analysis may reflect the presence of pleiotropy, which we were unable to directly test for in several of our analyses. We were able to test this for our MR of schizophrenia on cEF and found no evidence of pleiotropy. However, this should be considered when interpreting our other results.

### Limitations

There are a number of limitations to our study that should be considered when interpreting these results. First, there could be low statistical power to detect causal effects for some of the analyses. In particular, where anxiety, alcohol dependence and CUD are the exposures there were a low number of genome-wide significant SNPs. To overcome this issue we lowered the p-value threshold for our MR analyses to 1×10^−5^, but this means that any interpretation of these results should be approached with caution and revisiting this causal relationship when larger GWAS are available would be valuable. Second, in the majority of our analyses (i.e., in both directions for schizophrenia, MDD, smoking initiation, drinks per week and CUD) evidence of heterogeneity was observed in the IVW estimates, which could suggest that horizontal pleiotropy is present (e.g., that independent pathways are responsible for the influence of SNPs on the exposure and outcome). Therefore, caution should be used when interpreting these results. However, we did test for violations of other MR assumptions using additional MR sensitivity analyses and the direction of the results were consistent with the direction of the main results. Despite this, in a number of our analyses we could not test for directional pleiotropy due to the I-squared estimate being too low.

Finally, MR is also subject to some general limitations^40^. For example, the ‘Winner’s curse’ can occur when the SNPs used are based on the discovery GWAS only (as opposed to combined discovery and replication results), meaning that SNP-trait effects may be overestimated. This can mean that the MR estimate is biased towards the null. Thus, the focus of our results is on the direction of effect as opposed to the size of any causal effects, although the latter may still be somewhat informative.

### Conclusion

Our findings suggest a bidirectional causal relationship between cEF and schizophrenia liability, where increased schizophrenia liability is associated with decreased cEF and vice versa, as well as causal effects of increased cEF on reduced MDD liability and CUD liability and decreased drinks per week, and a causal effect of smoking initiation on decreased cEF. These results require further study to better understand the mechanisms behind these causal effects. Future research would benefit from better powered GWAS for anxiety where a lack of power may explain why we did not detect any causal effects. Our results may inform prioritisation of experimental medicine studies (e.g., of interventions targeting EF) to improve the likelihood of successful translation and suggest that these interventions may be useful prior to the onset of schizophrenia and MDD in particular.

## Acknowledgements

We thank all the contributors to the consortia we have used GWAS results from in our analyses. We would like the thank the research participants and employees of 23andMe for making this work possible.

## Author contributions

Conceptualization: MRM, ZER; Methodology: SMIB, ZER; Formal Analysis: SMIB, ZER; Resources: MRM; Data Curation: ZER; Writing—Original Draft: SMIB, ZER; Writing—Review and Editing: SMIB, HMS, ASH, MRM, ZER; Supervision: MRM, ZER; Project Administration: MRM, ZER; Funding Acquisition: MRM.

## References

1. Snyder, H. R., Miyake, A. & Hankin, B. L. Advancing understanding of executive function impairments and psychopathology: Bridging the gap between clinical and cognitive approaches. Front. Psychol. 6, 328 (2015).

2. Kim-Spoon, J. et al. Executive functioning and substance use in adolescence: Neurobiological and behavioral perspectives. Neuropsychologia 100, 79–92 (2017).

3. Thai, M. L., Andreassen, A. K. & Bliksted, V. A meta-analysis of executive dysfunction in patients with schizophrenia: Different degree of impairment in the ecological subdomains of the Behavioural Assessment of the Dysexecutive Syndrome. Psychiatry Research vol. 272 230–236 (2019).

4. Fioravanti, M., Bianchi, V. & Cinti, M. E. Cognitive deficits in schizophrenia: An updated metanalysis of the scientific evidence. BMC Psychiatry 12, 64 (2012).

5. Thuaire, F. et al. Executive functions in schizophrenia aging: Differential effects of age within specific executive functions. Cortex 125, 109–121 (2020).

6. Rock, P. L., Roiser, J. P., Riedel, W. J. & Blackwell, A. D. Cognitive impairment in depression: a systematic review and meta-analysis. Psychol. Med. 44, 2029–40 (2014).

7. Schwert, C., Stohrer, M., Aschenbrenner, S., Weisbrod, M. & Schröder, A. Neurocognitive profile of outpatients with unipolar depressive disorders. J. Clin. Exp. Neuropsychol. 41, 913–924 (2019).

8. Gulpers, B., Lugtenburg, A., Zuidersma, M., Verhey, F. R. J. & Voshaar, R. C. O. Anxiety disorders and figural fluency: A measure of executive function. J. Affect. Disord. 234, 38–44 (2018).

9. Murphy, Y. E. et al. An Investigation of Executive Functioning in Pediatric Anxiety. Behav. Modif. 42, 885–913 (2018).

10. Glass, J. M. et al. Effects of alcoholism severity and smoking on executive neurocognitive function. Addiction 104, 38–48 (2009).

11. Heffernan, T. M., Carling, A., O’Neill, T. S. & Hamilton, C. Smoking impedes executive function and related prospective memory. Ir. J. Psychol. Med. 31, 159–165 (2014).

12. Lundqvist, T. Cognitive consequences of cannabis use: Comparison with abuse of stimulants and heroin with regard to attention, memory and executive functions. Pharmacol. Biochem. Behav. 81, 319–330 (2005).

13. Lorenzetti, V., Hoch, E. & Hall, W. Adolescent cannabis use, cognition, brain health and educational outcomes: A review of the evidence. Eur. Neuropsychopharmacol. 36, 169–180 (2020).

14. Martins, J. S., Bartholow, B. D., Cooper, M. L., Von Gunten, C. D. & Wood, P. K. Associations between executive functioning, affect-regulation drinking motives, and alcohol use and problems. Psychol. Addict. Behav. 32, 16–28 (2018).

15. Diamond, A. & Ling, D. S. Conclusions about interventions, programs, and approaches for improving executive functions that appear justified and those that, despite much hype, do not. Dev. Cogn. Neurosci. 18, 34–48 (2016).

16. Hatoum, A. S. et al. Genome-Wide Association Study of Over 427,000 Individuals Establishes Executive Functioning as a Neurocognitive Basis of Psychiatric Disorders Influenced by GABAergic Processes. bioRxiv 674515 (2020) doi:10.1101/674515.

17. Davey Smith, G. & Ebrahim, S. ‘Mendelian randomization’: Can genetic epidemiology contribute to understanding environmental determinants of disease? International Journal of Epidemiology vol. 32 1–22 (2003).

18. Gupta, V., Walia, G. K. & Sachdeva, M. P. ‘Mendelian randomization’: an approach for exploring causal relations in epidemiology. Public Health vol. 145 113–119 (2017).

19. Carnegie, R. et al. Mendelian randomisation for nutritional psychiatry. The lancet. Psychiatry 7, 208–216 (2020).

20. Miyake, A. et al. The Unity and Diversity of Executive Functions and Their Contributions to Complex ‘Frontal Lobe’ Tasks: A Latent Variable Analysis. Cogn. Psychol. 41, 49–100 (2000).

21. Schizophrenia Working Group of the Psychiatric Genomics Consortium., Ripke, S., Walters, J. T. & O’Donovan, M. C. Mapping genomic loci prioritises genes and implicates synaptic biology in schizophrenia. medRxiv 2020.09.12.20192922 (2020) doi:10.1101/2020.09.12.20192922.

22. Wray, N. R. et al. Genome-wide association analyses identify 44 risk variants and refine the genetic architecture of major depression. Nat. Genet. 50, 668–681 (2018).

23. Otowa, T. et al. Meta-analysis of genome-wide association studies of anxiety disorders. Mol. Psychiatry 21, 1391–1399 (2016).

24. Liu, M. et al. Association studies of up to 1.2 million individuals yield new insights into the genetic etiology of tobacco and alcohol use. Nature Genetics vol. 51 237–244 (2019).

25. Walters, R. K. et al. Transancestral GWAS of alcohol dependence reveals common genetic underpinnings with psychiatric disorders. Nat. Neurosci. 21, 1656–1669 (2018).

26. Johnson, E. C. et al. A large-scale genome-wide association study meta-analysis of cannabis use disorder. The Lancet Psychiatry 7, 1032–1045 (2020).

27. Mägi, R. & Morris, A. P. GWAMA: Software for genome-wide association meta-analysis. BMC Bioinformatics 11, 288 (2010).

28. R Core Team. R: A language and environment for statistical computing. (2016).

29. Hemani, G., Tilling, K. & Davey Smith, G. Orienting the causal relationship between imprecisely measured traits using GWAS summary data. PLOS Genet. 13, e1007081 (2017).

30. Hemani, G. et al. The MR-base platform supports systematic causal inference across the human phenome. Elife 7, 1–29 (2018).

31. Machiela, M. J. & Chanock, S. J. LDlink: A web-based application for exploring population-specific haplotype structure and linking correlated alleles of possible functional variants. Bioinformatics 31, 3555–3557 (2015).

32. Burgess, S., Butterworth, A. & Thompson, S. G. Mendelian randomization analysis with multiple genetic variants using summarized data. Genet. Epidemiol. 37, 658–665 (2013).

33. Bowden, J., Davey Smith, G. & Burgess, S. Mendelian randomization with invalid instruments: Effect estimation and bias detection through Egger regression. Int. J. Epidemiol. 44, 512–525 (2015).

34. Bowden, J., Davey Smith, G., Haycock, P. C. & Burgess, S. Consistent Estimation in Mendelian Randomization with Some Invalid Instruments Using a Weighted Median Estimator. Genet. Epidemiol. 40, 304–314 (2016).

35. Hartwig, F. P., Davey Smith, G. & Bowden, J. Robust inference in summary data Mendelian randomization via the zero modal pleiotropy assumption. Int. J. Epidemiol. (2017) doi:10.1093/ije/dyx102.

36. Hemani, G., Tilling, K. & Davey Smith, G. Orienting the causal relationship between imprecisely measured traits using GWAS summary data. PLoS Genet. 13, e1007081 (2017).

37. Bowden, J. et al. Assessing the suitability of summary data for two-sample mendelian randomization analyses using MR-Egger regression: The role of the I 2 statistic. Int. J. Epidemiol. 45, 1961–1974 (2016).

38. Hettema, J. M. What is the genetic relationship between anxiety and depression? Am. J. Med. Genet. Part C Semin. Med. Genet. 148C, 140–146 (2008).

39. Hines, L. A., Treur, J. L., Jones, H. J., Sallis, H. M. & Munafò, M. R. Using genetic information to inform policy on cannabis. The Lancet Psychiatry 7, 1002–1003 (2020).

40. Haycock, P. C. et al. Best (but oft-forgotten) practices: The design, analysis, and interpretation of Mendelian randomization studies. Am. J. Clin. Nutr. 103, 965–978 (2016).

