## Supplementary materials for "Is there a causal relationship between executive function and liability to mental health and substance use? A Mendelian randomisation approach"

**Supplementary Table S1.** Weighted and unweighted I-squared results for all Mendelian randomisation analyses.

| **Exposure** | **Outcome** | **Mean F Statistic** | **Unweighted I-squared** | **Weighted I-squared** |
| --- | --- | --- | --- | --- |
| cEF | Schizophrenia | 41.85 | 0.47 | 0.26 |
| cEF | MDD | 41.74 | 0.46 | 0.12 |
| cEF | Anxiety | 41.87 | 0.44 | 0.86 |
| cEF | Smoking initiation | 41.74 | 0.46 | 0.38 |
| cEF | Drinks per week | 41.74 | 0.46 | 0.09 |
| cEF | Cannabis use disorder | 41.74 | 0.46 | 0.37 |
| cEF | Alcohol dependence | 41.74 | 0.46 | 0.45 |
| Schizophrenia | cEF | 45.11 | 0.60 | 0.32 |
| MDD | cEF | 11.83 | 0.20 | 0.07 |
| Anxiety | cEF | 21.75 | 0.42 | 0.00 |
| Smoking initiation | cEF | 31.01 | 0.59 | 0.51 |
| Drinks per week | cEF | 39.98 | 0.94 | 0.91 |
| Cannabis use disorder | cEF | 23.40 | 0.52 | 0.00 |
| Alcohol dependence | cEF | 23.32 | 0.76 | 0.00 |

*cEF=common executive function score, MDD=Major depressive disorder*

**Supplementary Table S2.** Bi-dirctional two-sample Mendelian randomisation results of executive function (cEF) on schizophrenia and MDD following Steiger filtering.

| Exposure | Outcome | Method | NSNPs | OR (95% CI) | P-value |
| --- | --- | --- | --- | --- | --- |
| cEF | Schizophrenia | IVW | 37/90 (41%) | 0.52 (0.35, 0.76) | 7.35x10^-04^ |
|  |  | MR Egger |  | 0.61 (0.10, 3.56) | 0.59 |
|  |  | Weighted median |  | 0.60 (0.34, 1.03) | 0.07 |
|  |  | Simple mode |  | 0.18 (0.04, 0.72) | 0.02 |
|  |  | Weighted mode |  | 0.18 (0.05, 0.63) | 0.01 |
| cEF | MDD | IVW | 74/91 (81%) | 0.68 (0.51, 0.90) | 6.87x10^-03^ |
|  |  | MR Egger |  | 0.38 (0.10, 1.48) | 0.17 |
|  |  | Weighted median |  | 0.55 (0.36, 0.84) | 5.56x10^-03^ |
|  |  | Simple mode |  | 0.33 (0.11, 1.00) | 0.05 |
|  |  | Weighted mode |  | 0.31 (0.12, 0.83) | 0.02 |
| Schizophrenia | cEF | - | 198/198 (100%) | - | - |
| MDD | cEF | - | 30/30 (100%) | - | - |

*With Steiger filtering, the variance each SNP explains in the exposure and outcome is calculated. The number of SNPs explaining more variance in the exposure than the outcome are presented in the NSNPs column. MR analyses were then repeated using only these SNPs. Where cEF was the outcome for schizophrenia and MDD, all SNPs included in the instruments for schizophrenia and MDD were better instruments for these than cEF and therefore these analyses was not repeated.*

*cEF=common executive function score, MDD=Major depressive disorder*

**Supplementary Figure S1.** Scatter plot for executive function as the exposure and schizophrenia as the outcome.

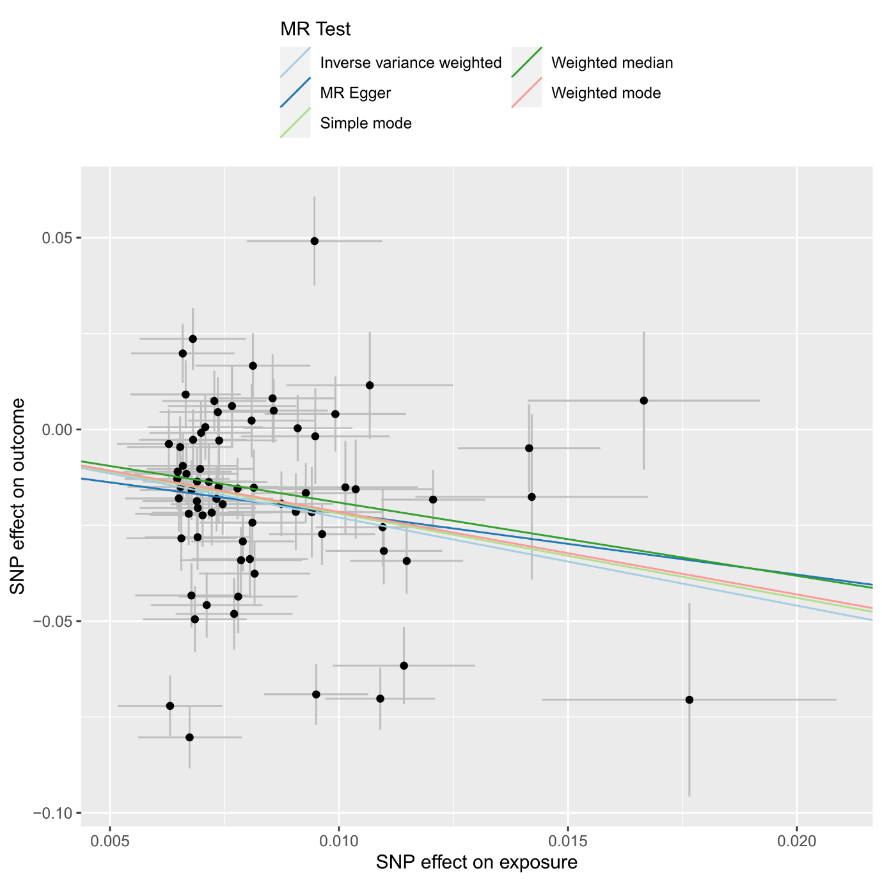

*Scatterplot showing the effect sizes of the SNP-exposure association and the SNP-outcome associations with standard error bars. The slopes of the lines correspond to causal estimates using each of the five different methods. All MR methods are in the same direction here.*

**Supplementary Figure S2.** Forest plot for executive function as the exposure and schizophrenia as the outcome.

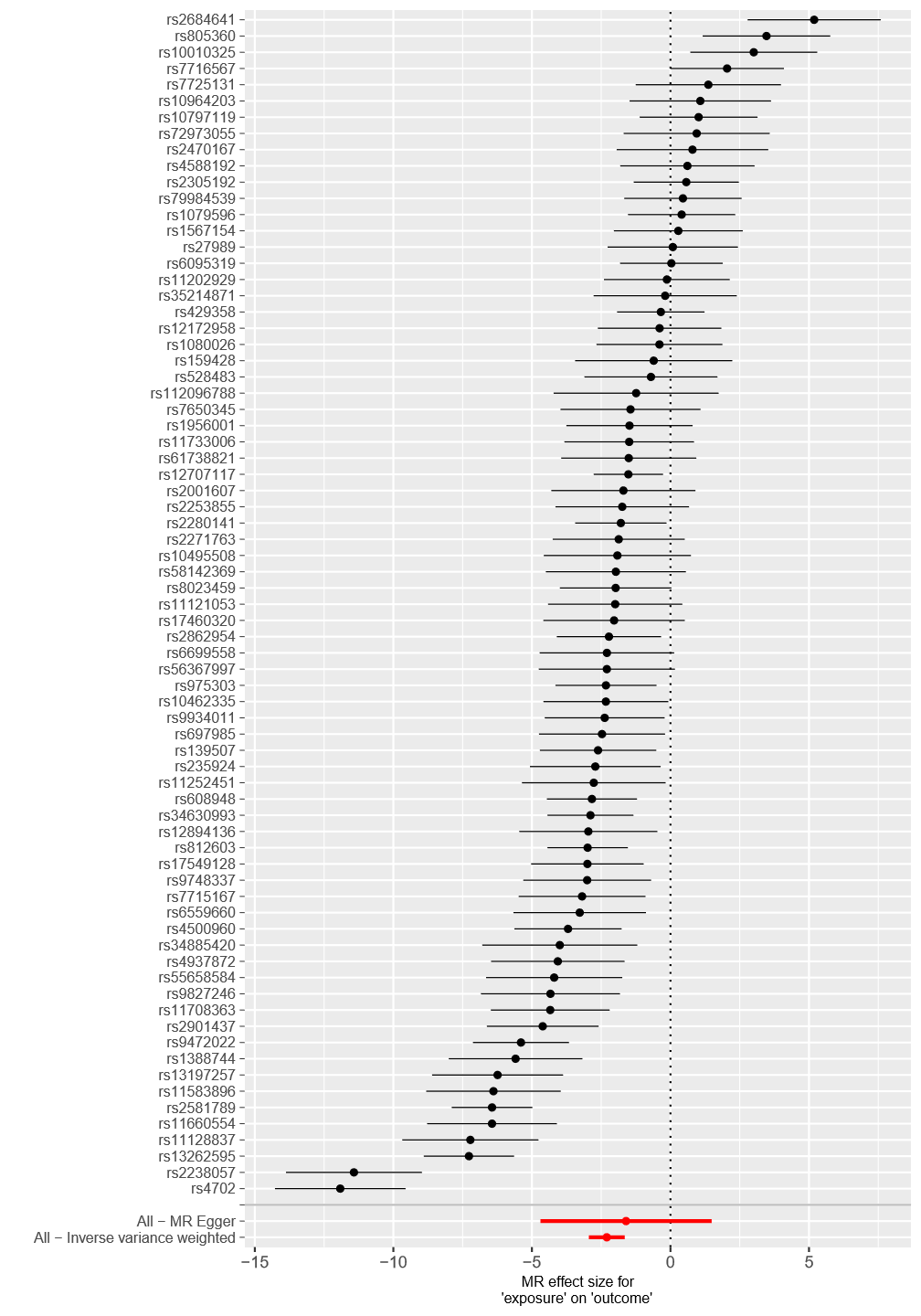

*Each black point represents the effect size for each SNP of the exposure on the outcome. Red points indicate the causal estimates for all SNPs as a single genetic instrument for the MR-Egger and the inverse-variance weighted (IVW) methods. Horizontal lines for each point represent 95% confidence intervals for each estimate. Some heterogeneity is observed here.*

**Supplementary Figure S3.** Funnel plot for executive function as the exposure and schizophrenia as the outcome.

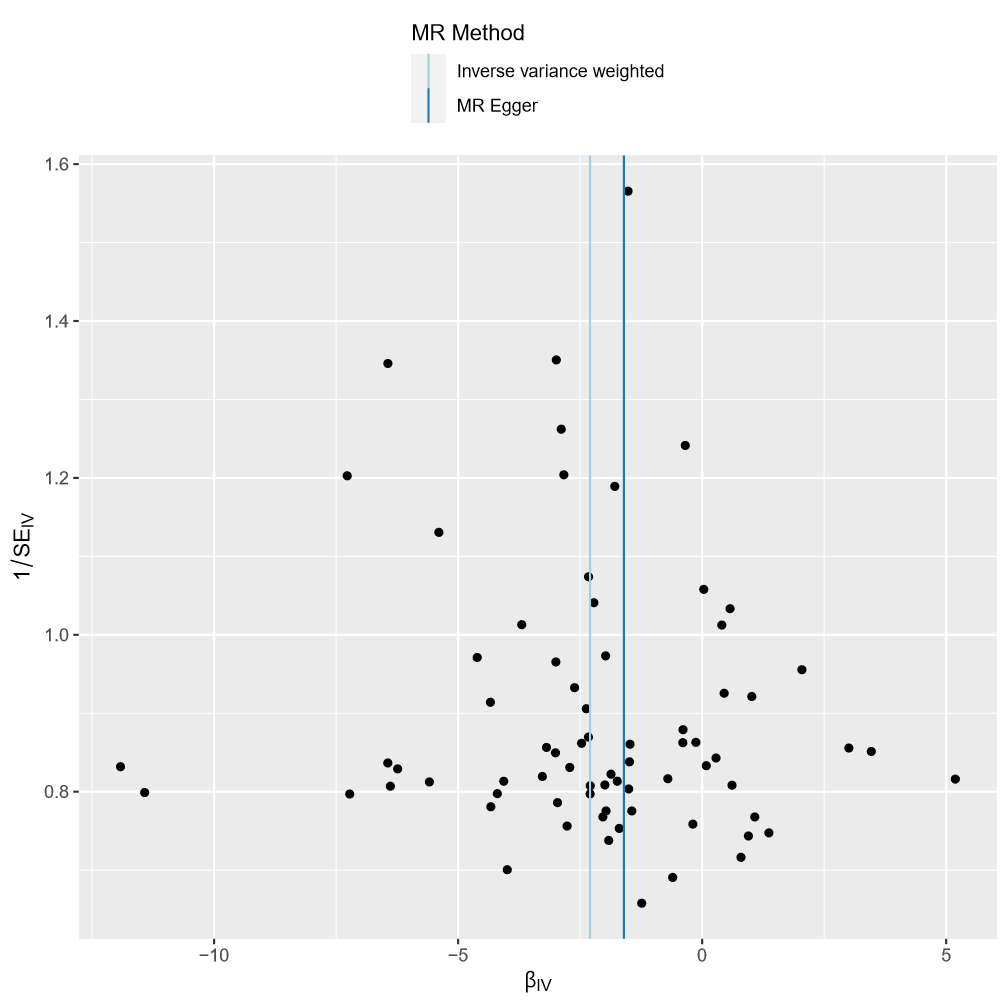

*Each SNP is used as a separate instrument to estimate a causal effect against the inverse of the standard error of that causal estimate. The vertical lines show the causal estimates for all SNPs using the IVW and MR-Egger methods. In this plot there is a degree of asymmetry, which may indicate violation of instrumental variable assumptions, e.g., horizontal pleiotropy.*

**Supplementary Figure S4.** Leave one out plot for executive function as the exposure and schizophrenia as the outcome.

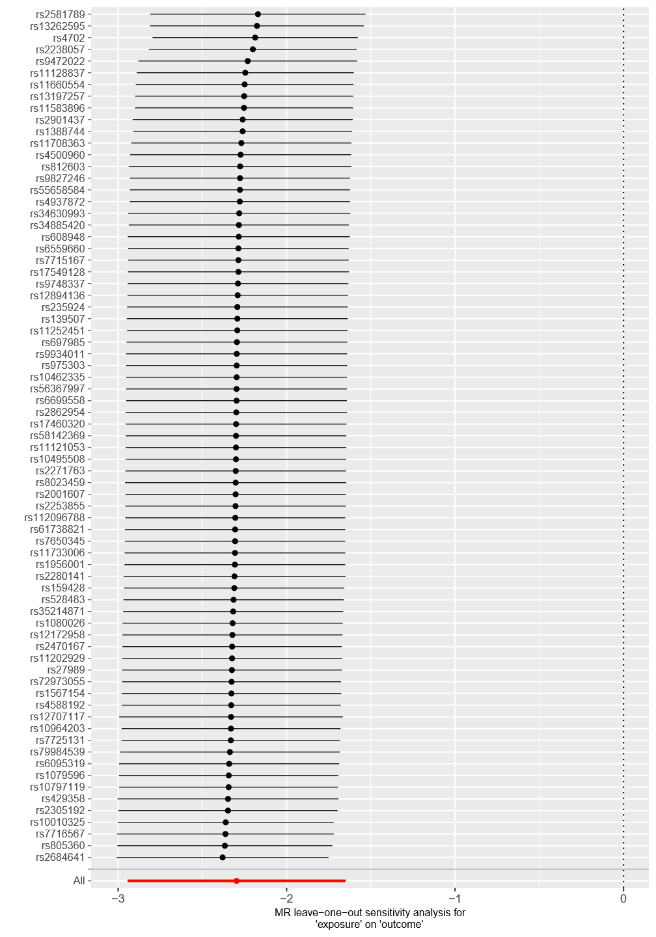

*Each black point represents the IVW estimate of the exposure on the outcome excluding that particular SNP from the analysis. The red point shows the IVW estimate using all SNPs in the instrument. There are no instances where the exclusion of one particular SNP leads to dramatic changes in the overall result.*

**Supplementary Figure S5.** Scatter plot for executive function as the exposure and major depressive disorder as the outcome.

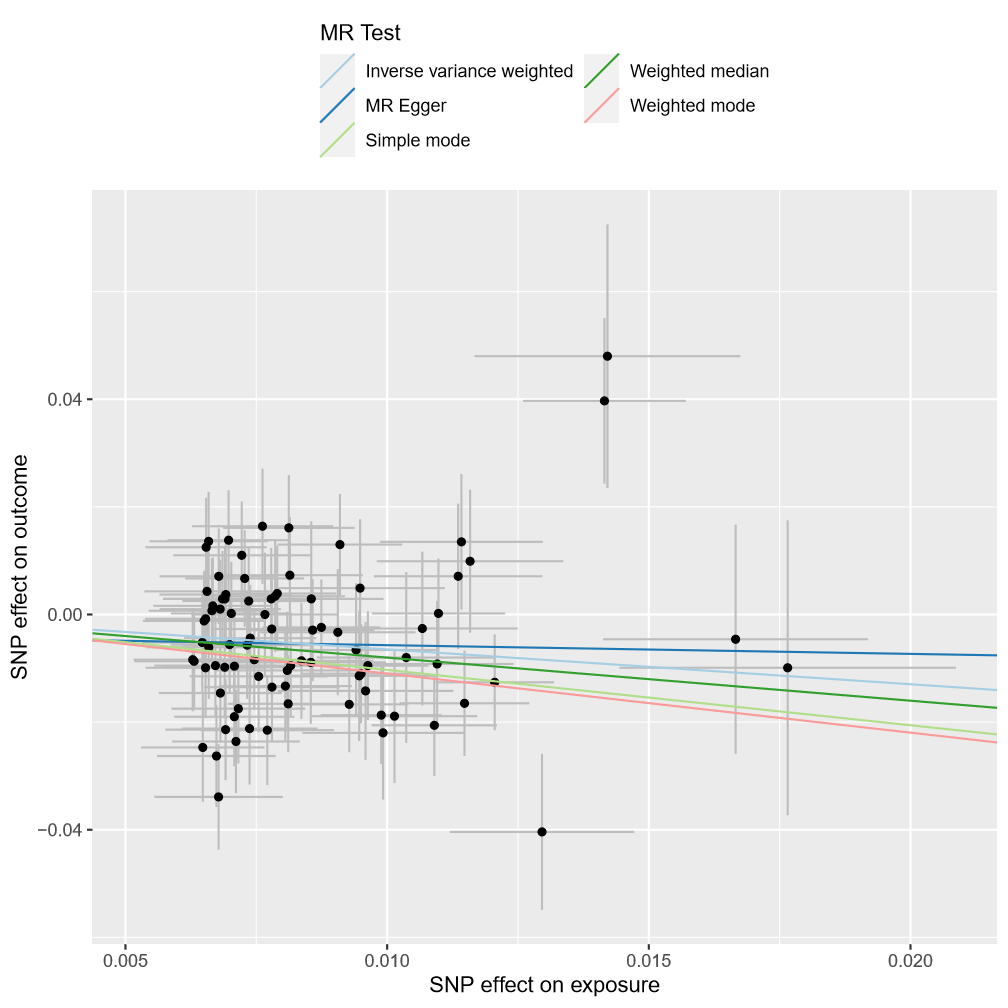

*Scatterplot showing the effect sizes of the SNP-exposure association and the SNP-outcome associations with standard error bars. The slopes of the lines correspond to causal estimates using each of the five different methods. All MR methods are in the same direction here.*

**Supplementary Figure S6.** Forest plot for executive function as the exposure and major depressive disorder as the outcome.

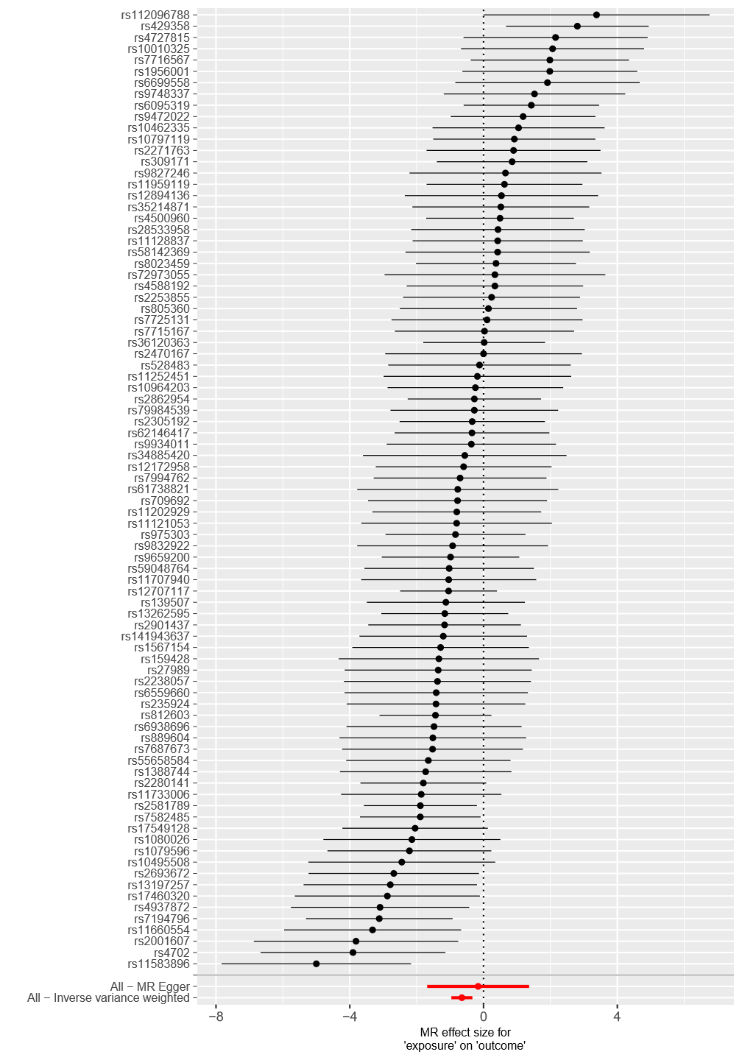

*Each black point represents the effect size for each SNP of the exposure on the outcome. Red points indicate the causal estimates for all SNPs as a single genetic instrument for the MR-Egger and the inverse-variance weighted (IVW) methods. Horizontal lines for each point represent 95% confidence intervals for each estimate. Some heterogeneity is observed here.*

**Supplementary Figure S7.** Funnel plot for executive function as the exposure and major depressive disorder as the outcome.

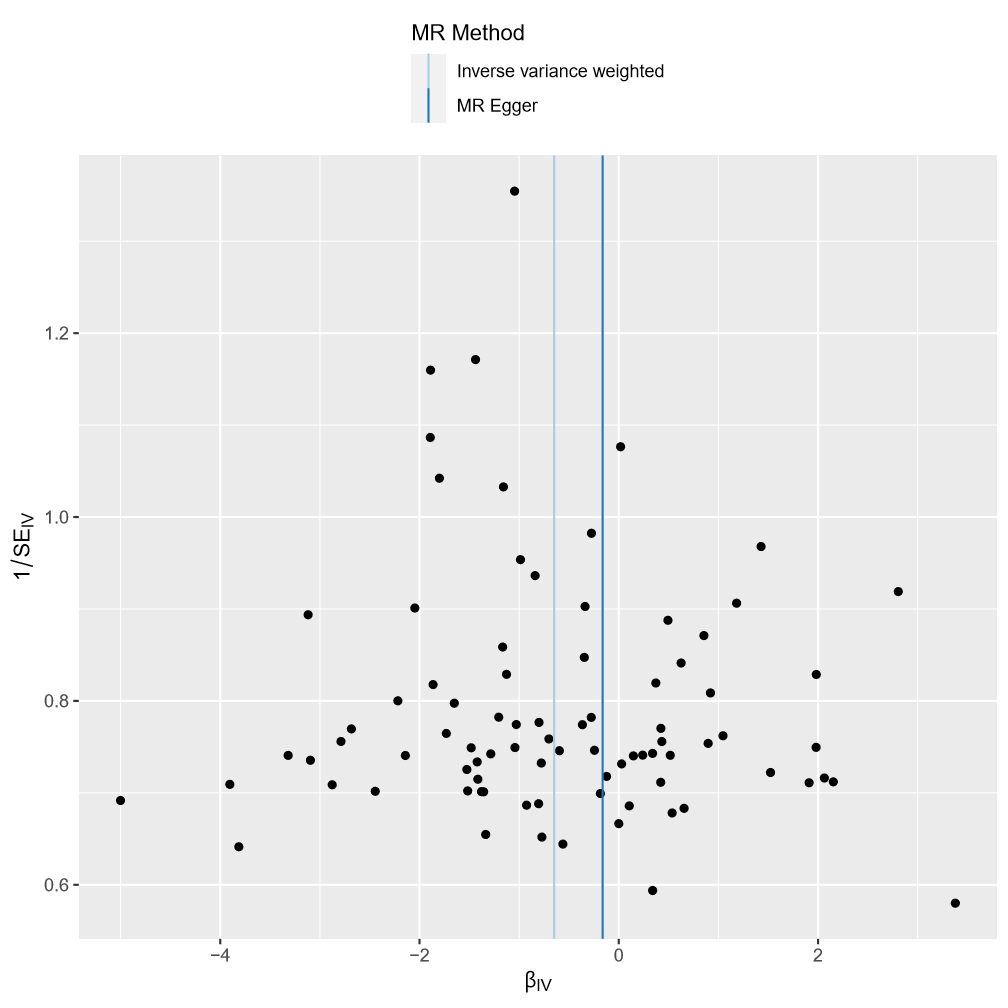

*Each SNP is used as a separate instrument to estimate a causal effect against the inverse of the standard error of that causal estimate. The vertical lines show the causal estimates for all SNPs using the IVW and MR-Egger methods. In this plot there is a degree of asymmetry, which may indicate violation of instrumental variable assumptions, e.g. horizontal pleiotropy.*
**Supplementary Figure S8.** Leave one out plot for executive function as the exposure and major depressive disorder as the outcome.

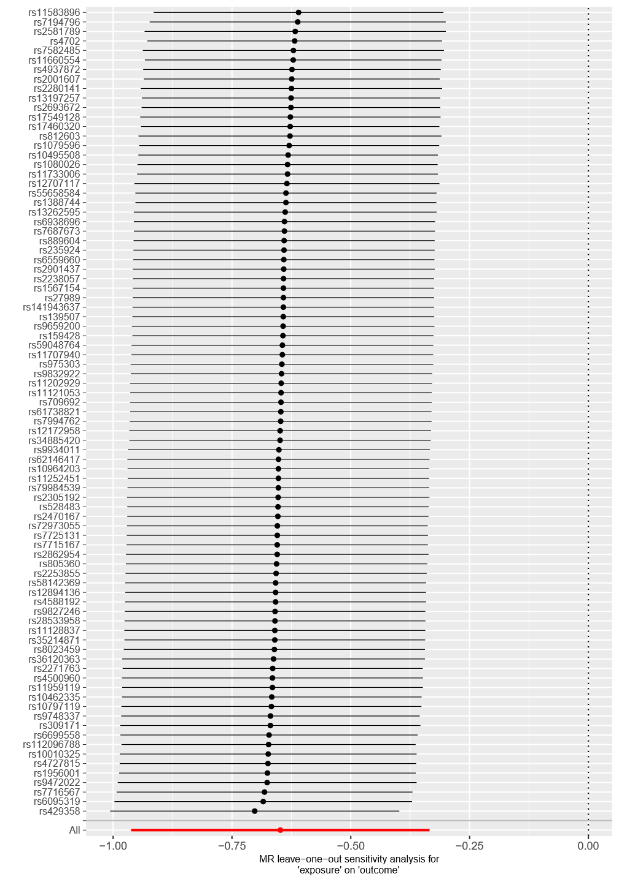

*Each black point represents the IVW estimate of the exposure on the outcome excluding that particular SNP from the analysis. The red point shows the IVW estimate using all SNPs in the instrument. There are no instances where the exclusion of one particular SNP leads to dramatic changes in the overall result.*

**Supplementary Figure S9.** Scatter plot for executive function as the exposure and drinks per week as the outcome.

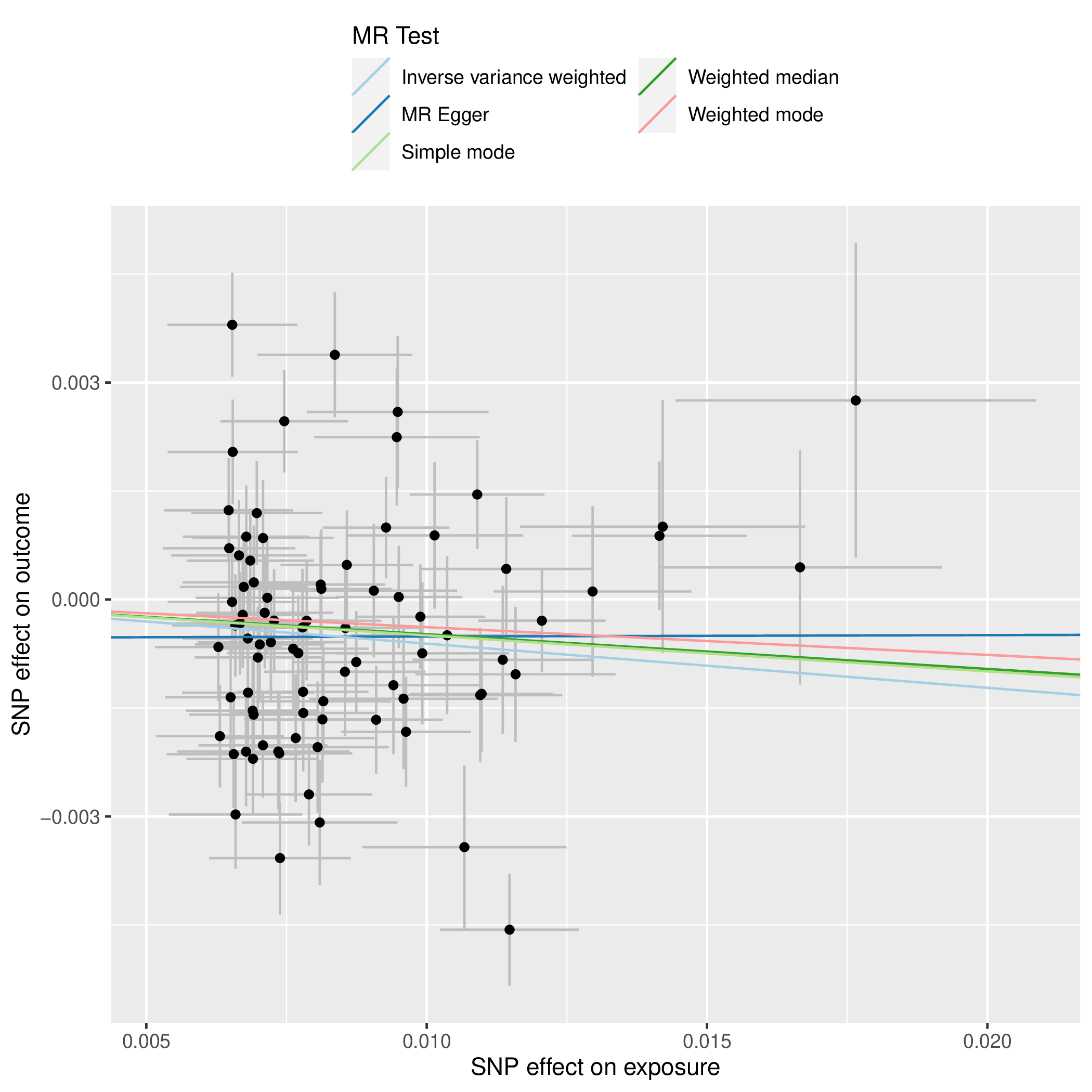

*Scatterplot showing the effect sizes of the SNP-exposure association and the SNP-outcome associations with standard error bars. The slopes of the lines correspond to causal estimates using each of the five different methods. All MR methods are in the same direction here.*

**Supplementary Figure S10.** Forest plot for executive function as the exposure and drinks per week as the outcome.

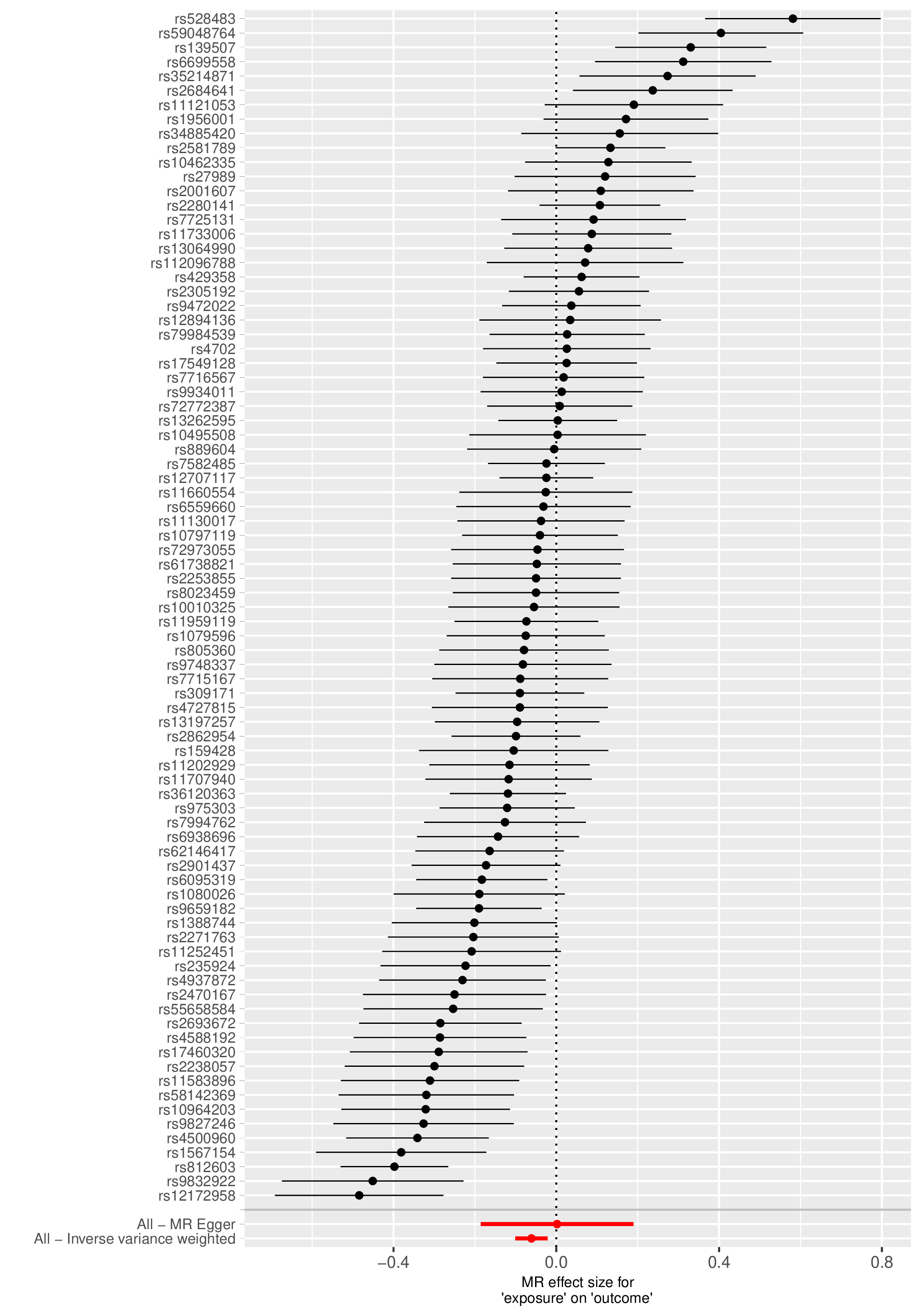

*Each black point represents the effect size for each SNP of the exposure on the outcome. Red points indicate the causal estimates for all SNPs as a single genetic instrument for the MR-Egger and the inverse-variance weighted (IVW) methods. Horizontal lines for each point represent 95% confidence intervals for each estimate. Some heterogeneity is observed here.*

**Supplementary Figure S11.** Funnel plot for executive function as the exposure and drinks per week as the outcome.

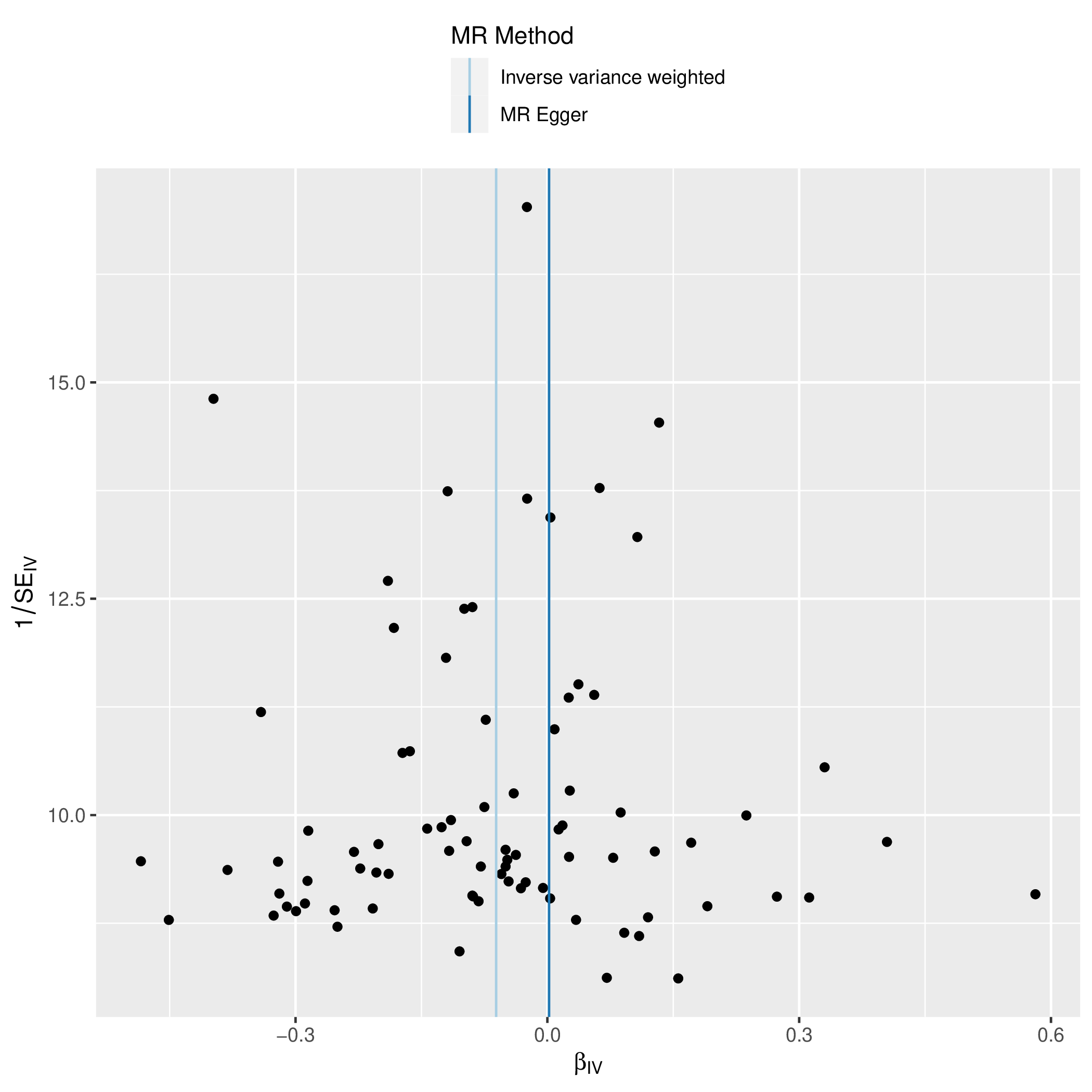

*Each SNP is used as a separate instrument to estimate a causal effect against the inverse of the standard error of that causal estimate. The vertical lines show the causal estimates for all SNPs using the IVW and MR-Egger methods. In this plot there is a degree of asymmetry, which may indicate violation of instrumental variable assumptions, e.g., horizontal pleiotropy.*

**Supplementary Figure S12.** Leave one out plot for executive function as the exposure and drinks per week as the outcome.

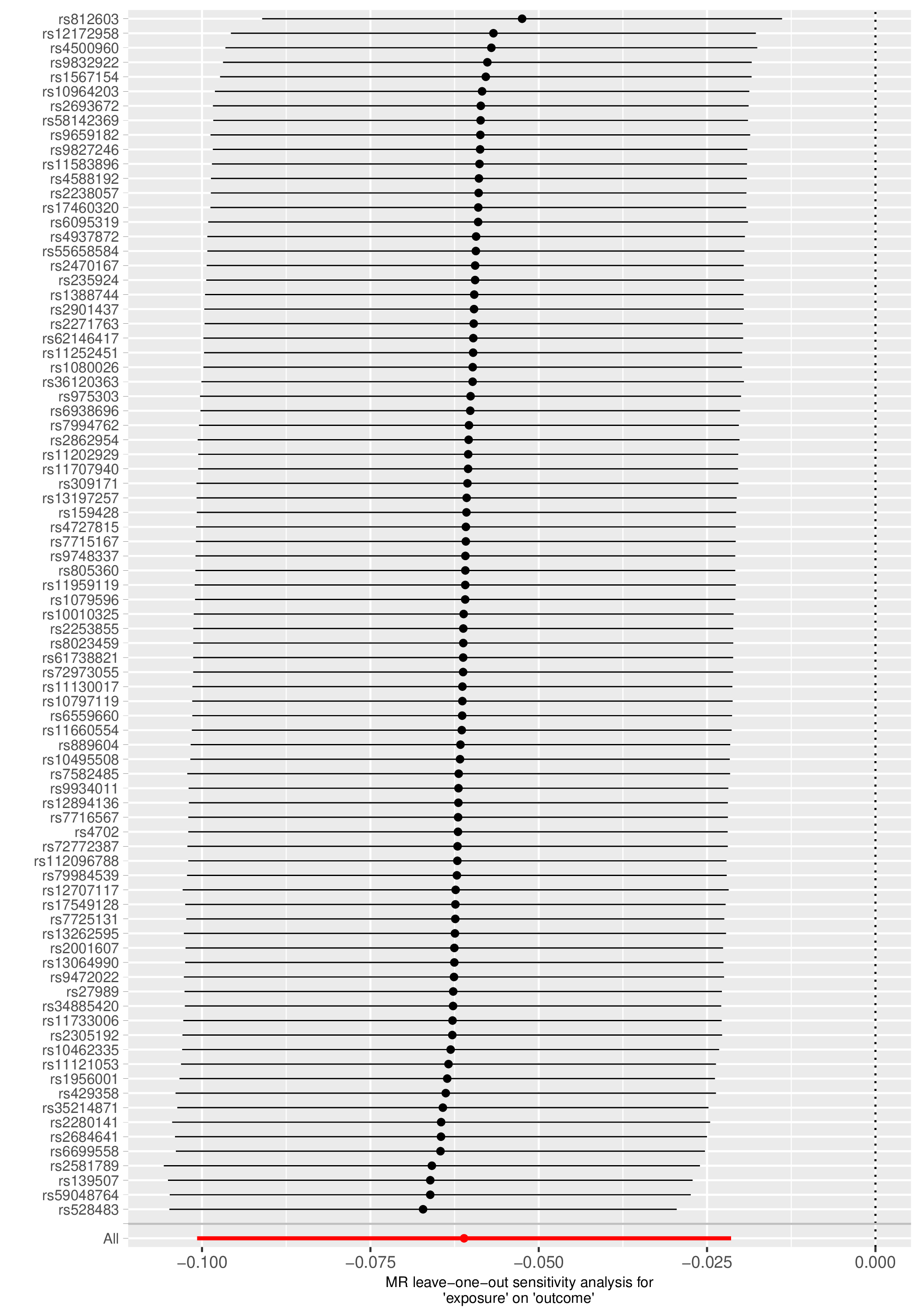

*Each black point represents the IVW estimate of the exposure on the outcome excluding that particular SNP from the analysis. The red point shows the IVW estimate using all SNPs in the instrument. There are no instances where the exclusion of one particular SNP leads to dramatic changes in the overall result.*

**Supplementary Figure S13.** Scatter plot for executive function as the exposure and cannabis use disorder as the outcome.

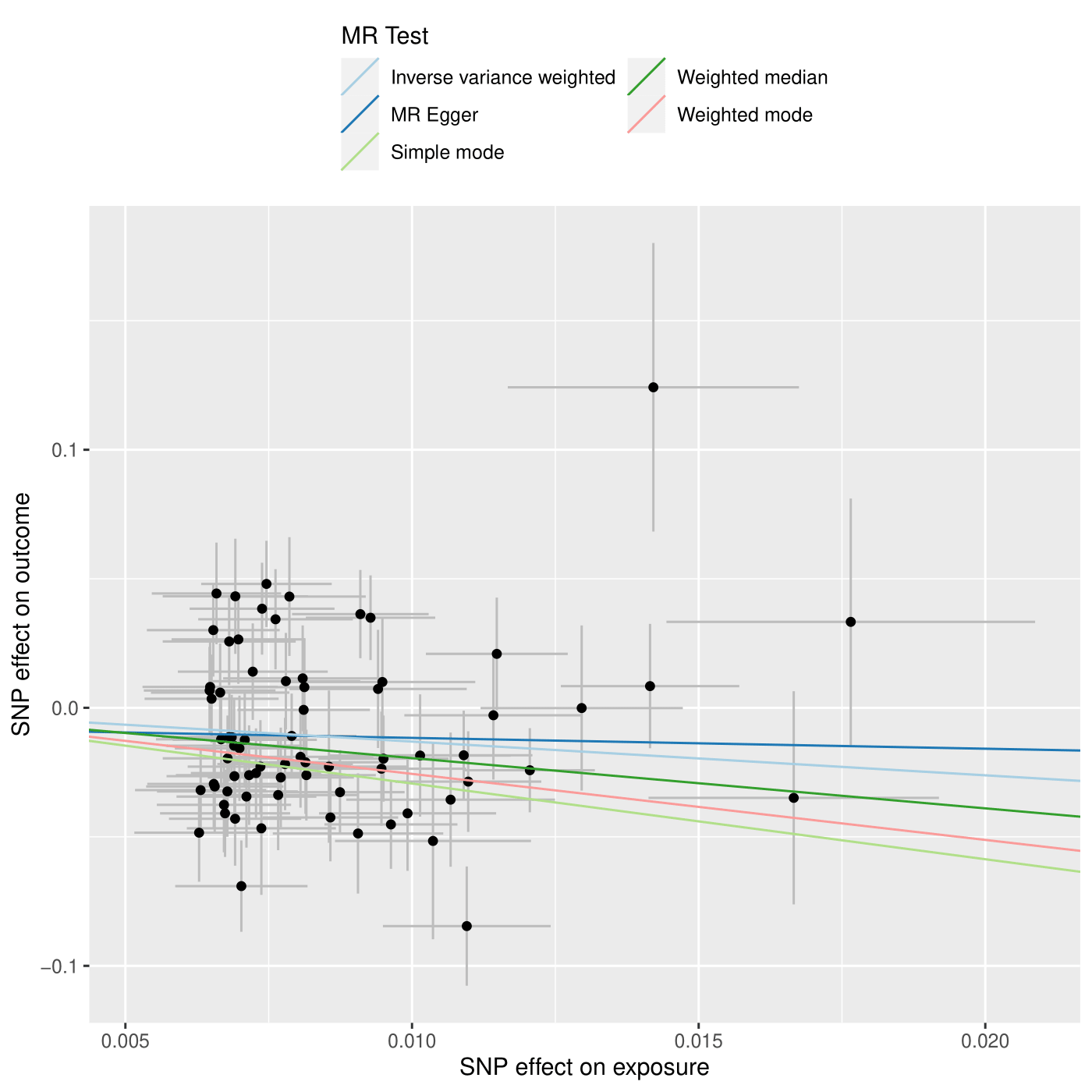

*Scatterplot showing the effect sizes of the SNP-exposure association and the SNP-outcome associations with standard error bars. The slopes of the lines correspond to causal estimates using each of the five different methods. All MR methods are in the same direction here.*

**Supplementary Figure S14.** Forest plot for executive function as the exposure and cannabis use disorder as the outcome.

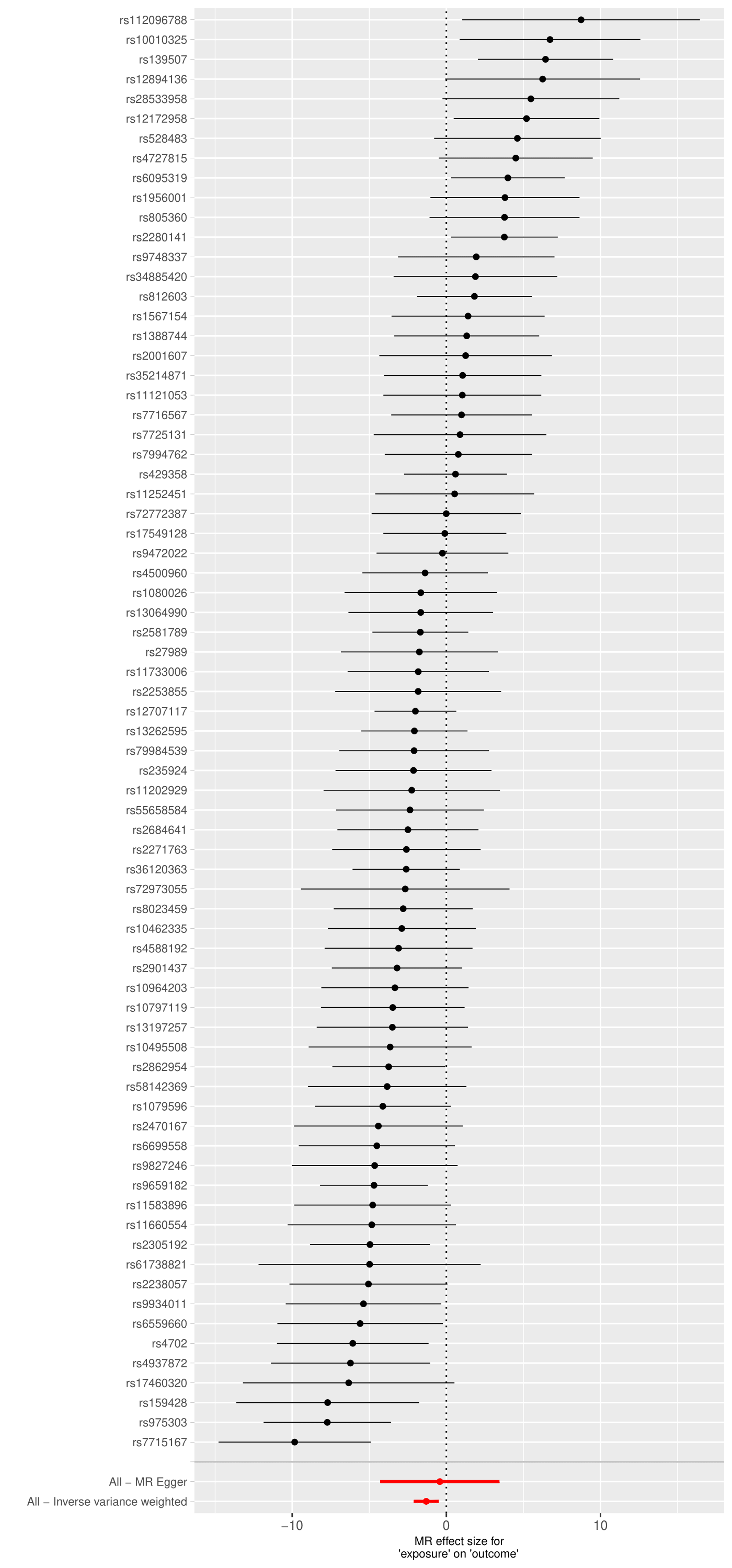

*Each black point represents the effect size for each SNP of the exposure on the outcome. Red points indicate the causal estimates for all SNPs as a single genetic instrument for the MR-Egger and the inverse-variance weighted (IVW) methods. Horizontal lines for each point represent 95% confidence intervals for each estimate. Some heterogeneity is observed here.*

**Supplementary Figure S15.** Funnel plot for executive function as the exposure and cannabis use disorder as the outcome.

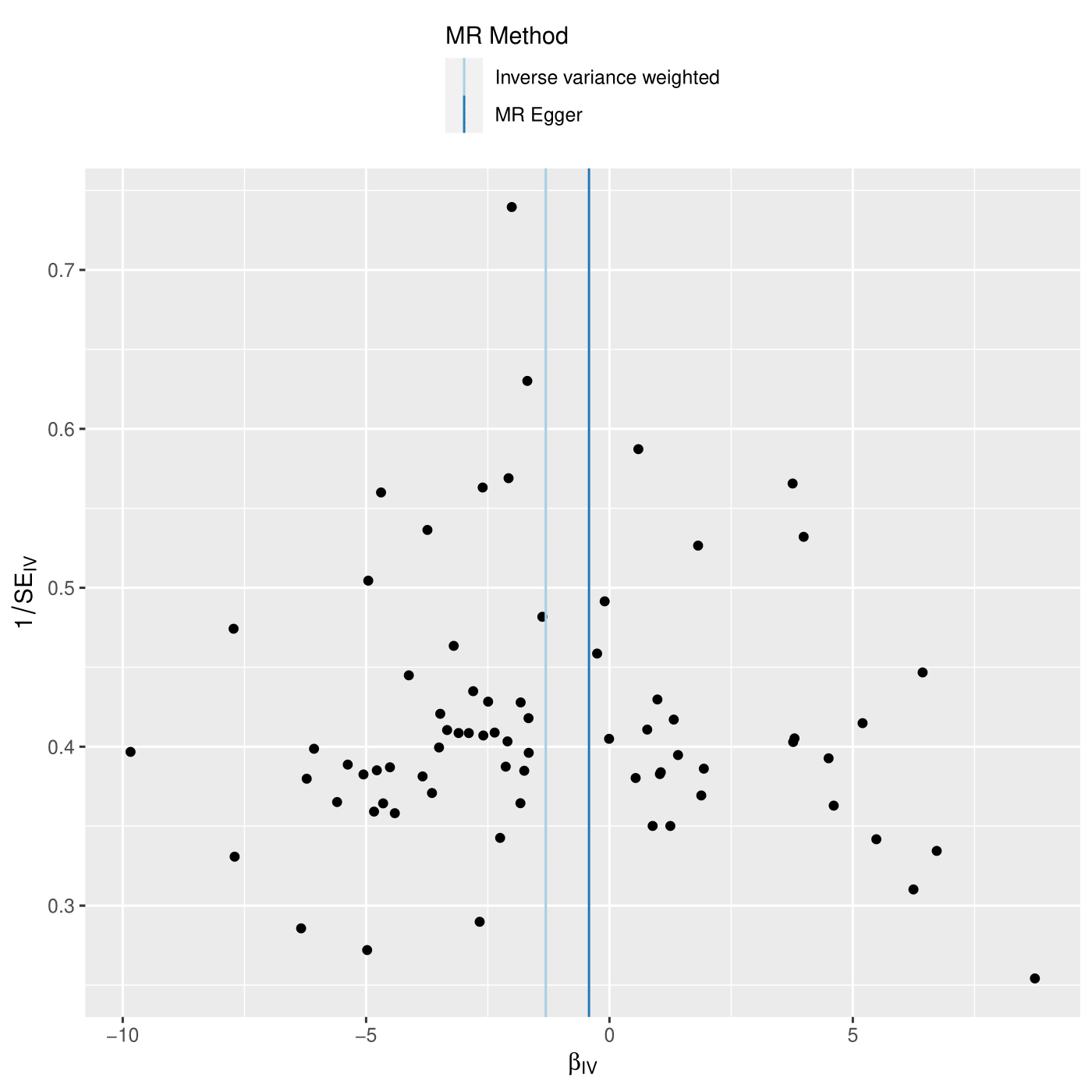

*Each SNP is used as a separate instrument to estimate a causal effect against the inverse of the standard error of that causal estimate. The vertical lines show the causal estimates for all SNPs using the IVW and MR-Egger methods. In this plot there slight asymmetry, which may indicate violation of instrumental variable assumptions, e.g., horizontal pleiotropy.*

**Supplementary Figure S16.** Leave one out plot for executive function as the exposure and cannabis use disorder as the outcome.

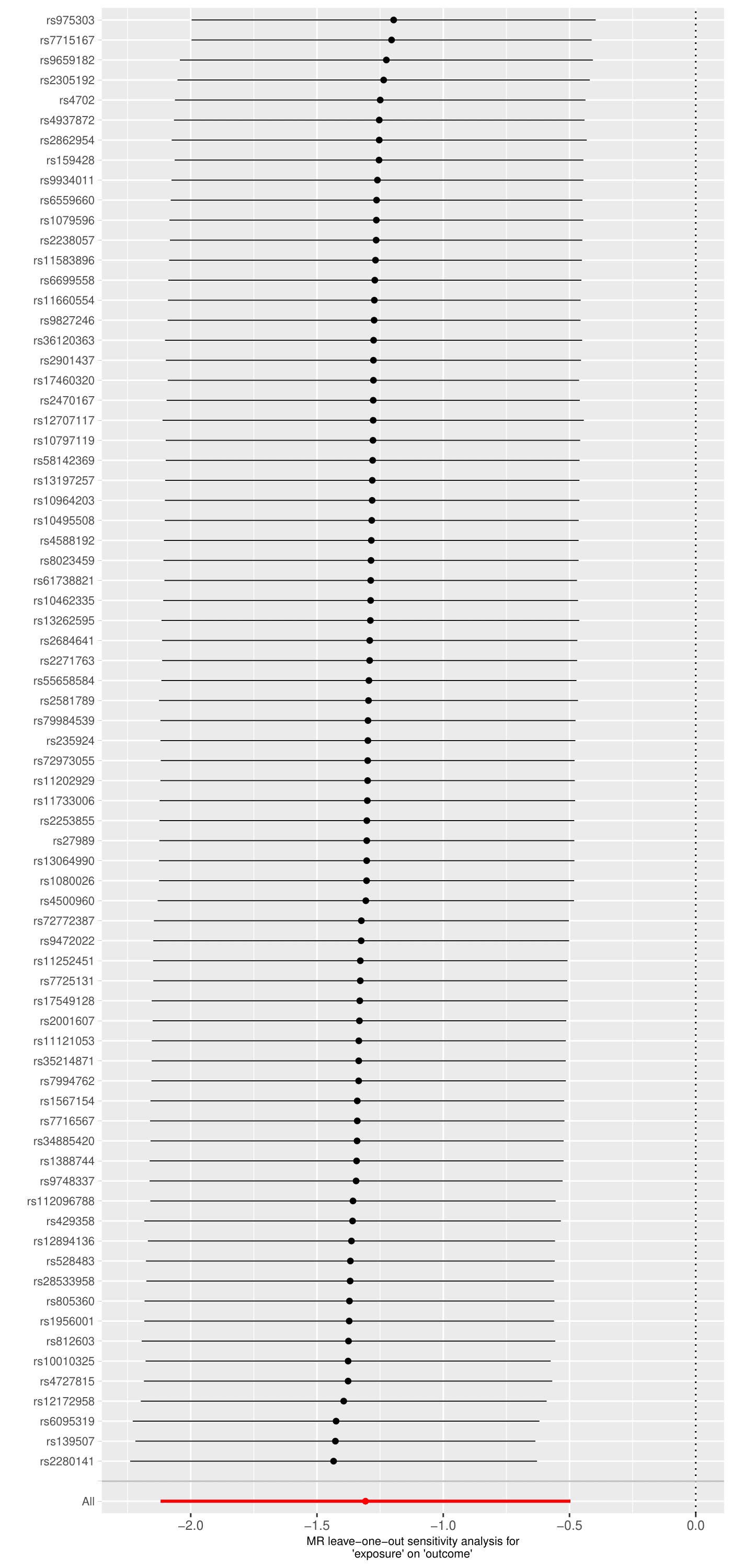

*Each black point represents the IVW estimate of the exposure on the outcome excluding that particular SNP from the analysis. The red point shows the IVW estimate using all SNPs in the instrument. There are no instances where the exclusion of one particular SNP leads to dramatic changes in the overall result.*

**Supplementary Figure S17.** Scatter plot for schizophrenia as the exposure and executive function as the outcome.

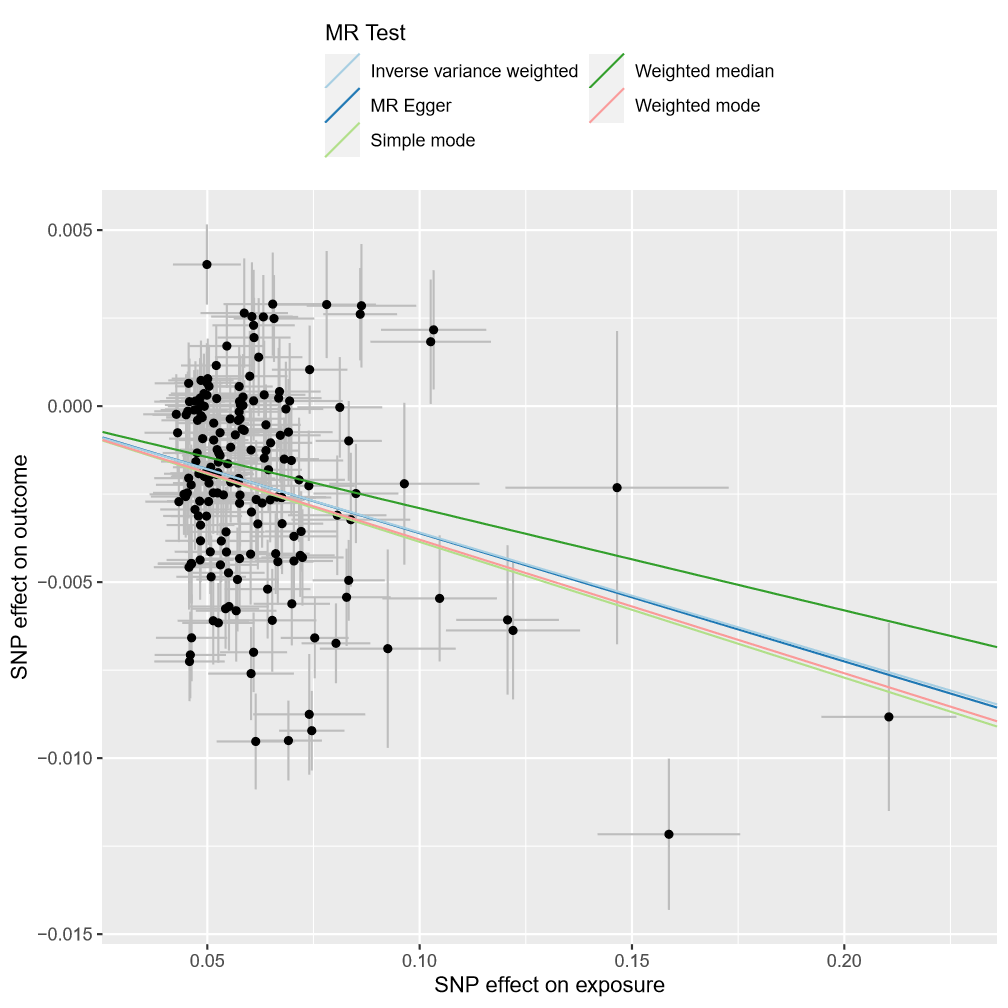

*Scatterplot showing the effect sizes of the SNP-exposure association and the SNP-outcome associations with standard error bars. The slopes of the lines correspond to causal estimates using each of the five different methods. All MR methods are in the same direction here.*

**Supplementary Figure S18.** Forest plot for schizophrenia as the exposure and executive function as the outcome.

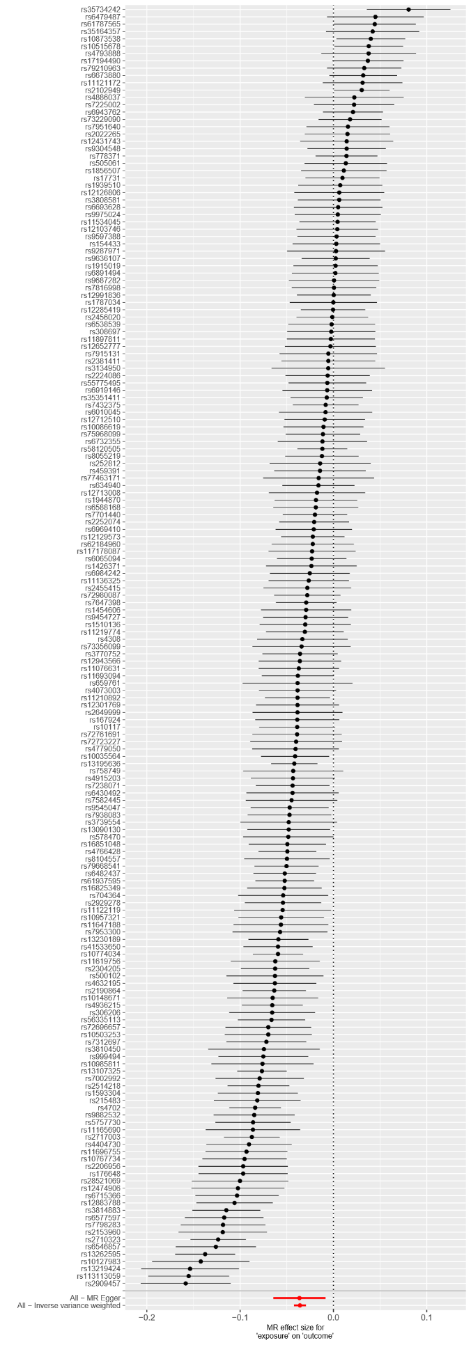

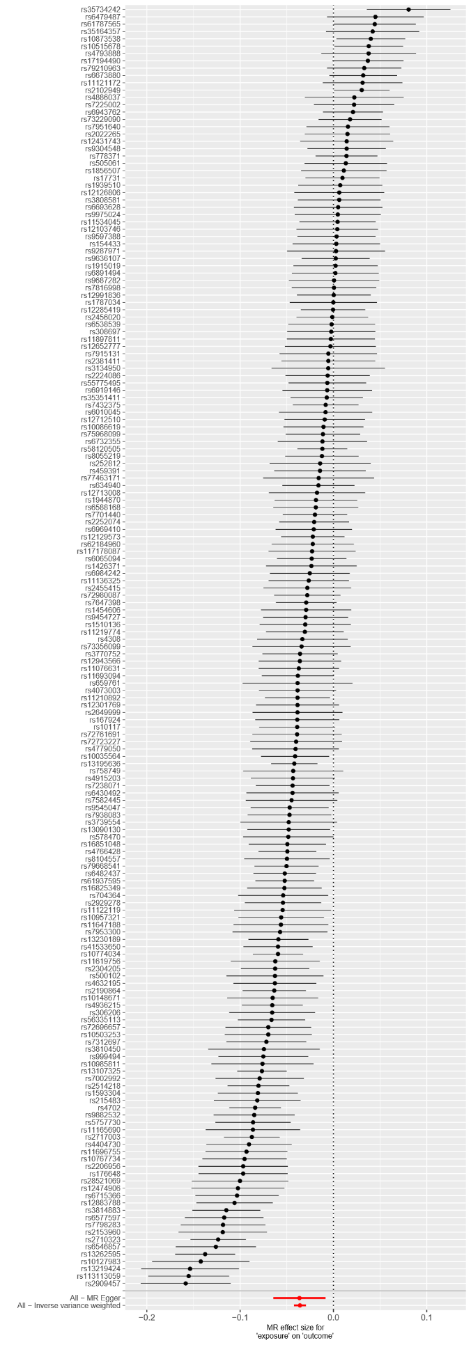

*Each black point represents the effect size for each SNP of the exposure on the outcome. Red points indicate the causal estimates for all SNPs as a single genetic instrument for the MR-Egger and the inverse-variance weighted (IVW) methods. Horizontal lines for each point represent 95% confidence intervals for each estimate. Some heterogeneity is observed here.*

**Supplementary Figure 19.** Funnel plot for schizophrenia as the exposure and executive function as the outcome.

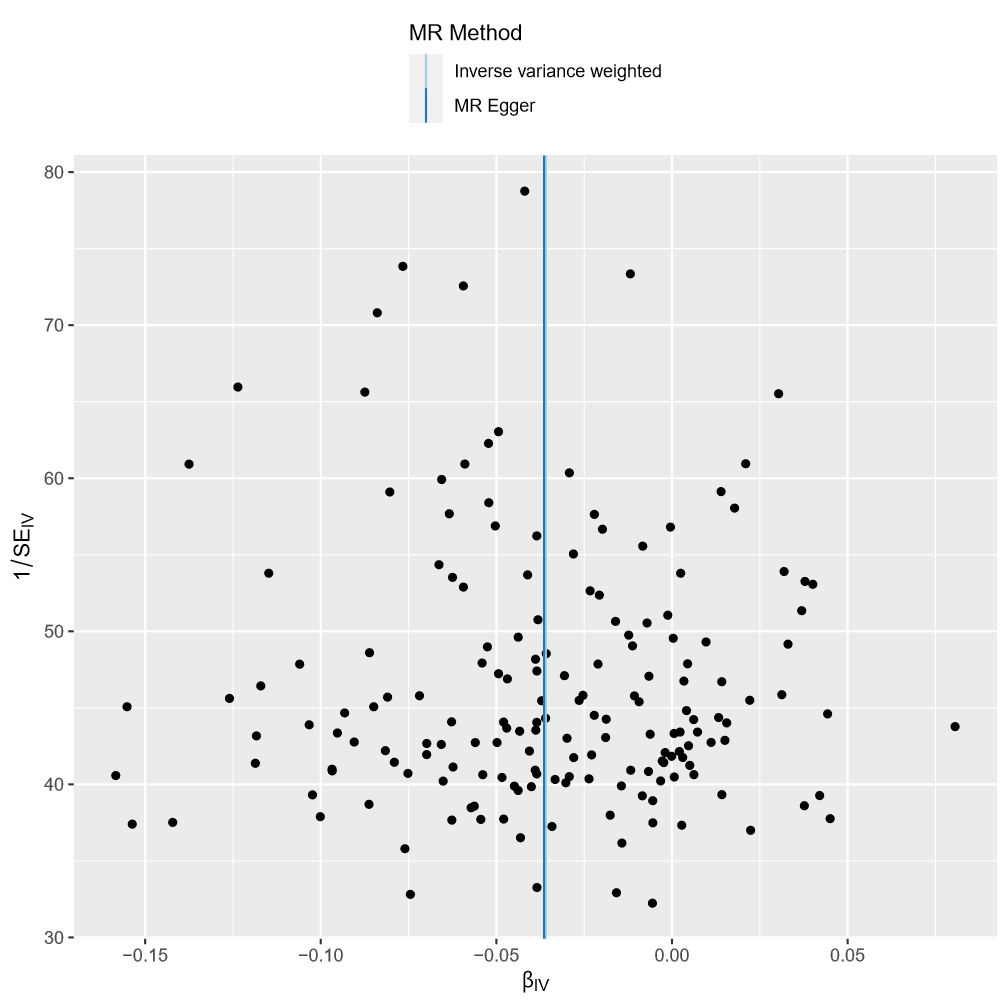

*Each SNP is used as a separate instrument to estimate a causal effect against the inverse of the standard error of that causal estimate. The vertical lines show the causal estimates for all SNPs using the IVW and MR-Egger methods. This plot is fairly symmetric.*

**Supplementary Figure S20.** Leave one out plot for schizophrenia as the exposure and executive function as the outcome.

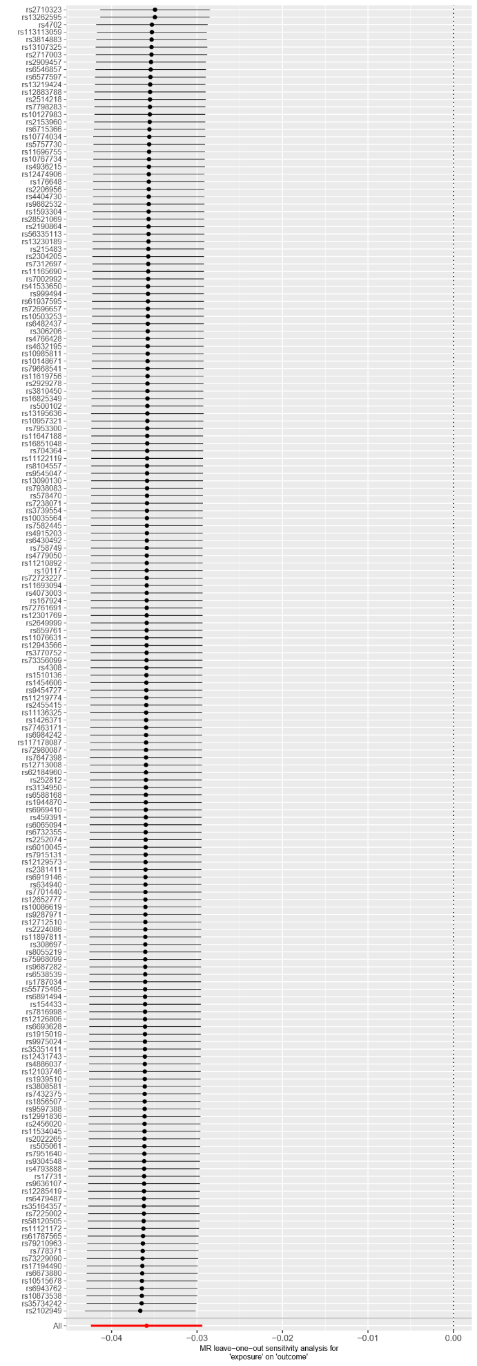

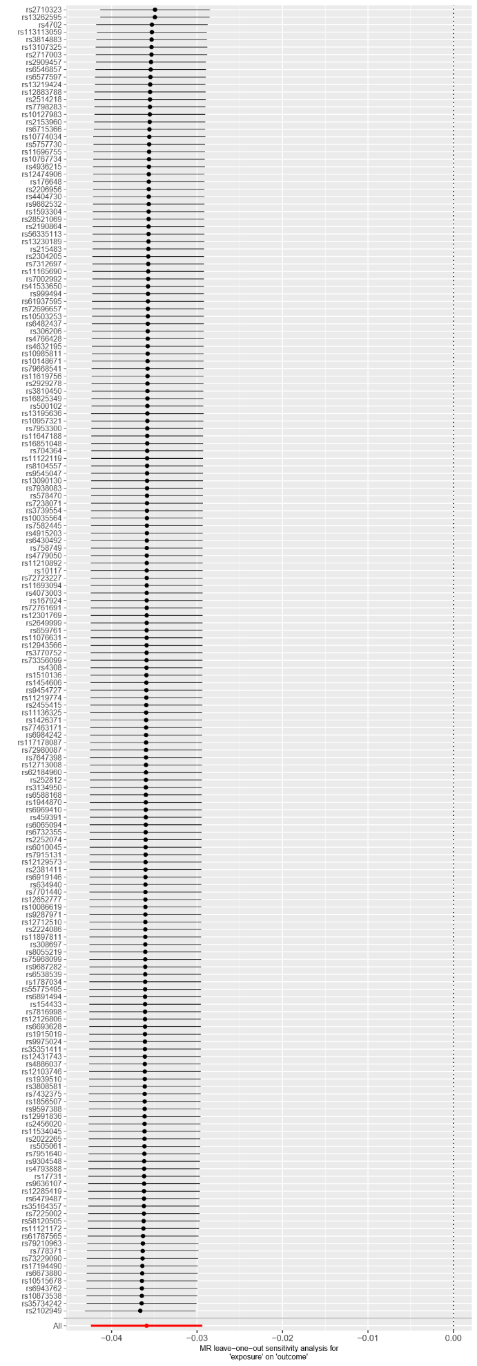

*Each black point represents the IVW estimate of the exposure on the outcome excluding that particular SNP from the analysis. The red point shows the IVW estimate using all SNPs in the instrument. There are no instances where the exclusion of one particular SNP leads to dramatic changes in the overall result.*

**Supplementary Figure S21.** Scatter plot for smoking initiation as the exposure and executive function as the outcome.

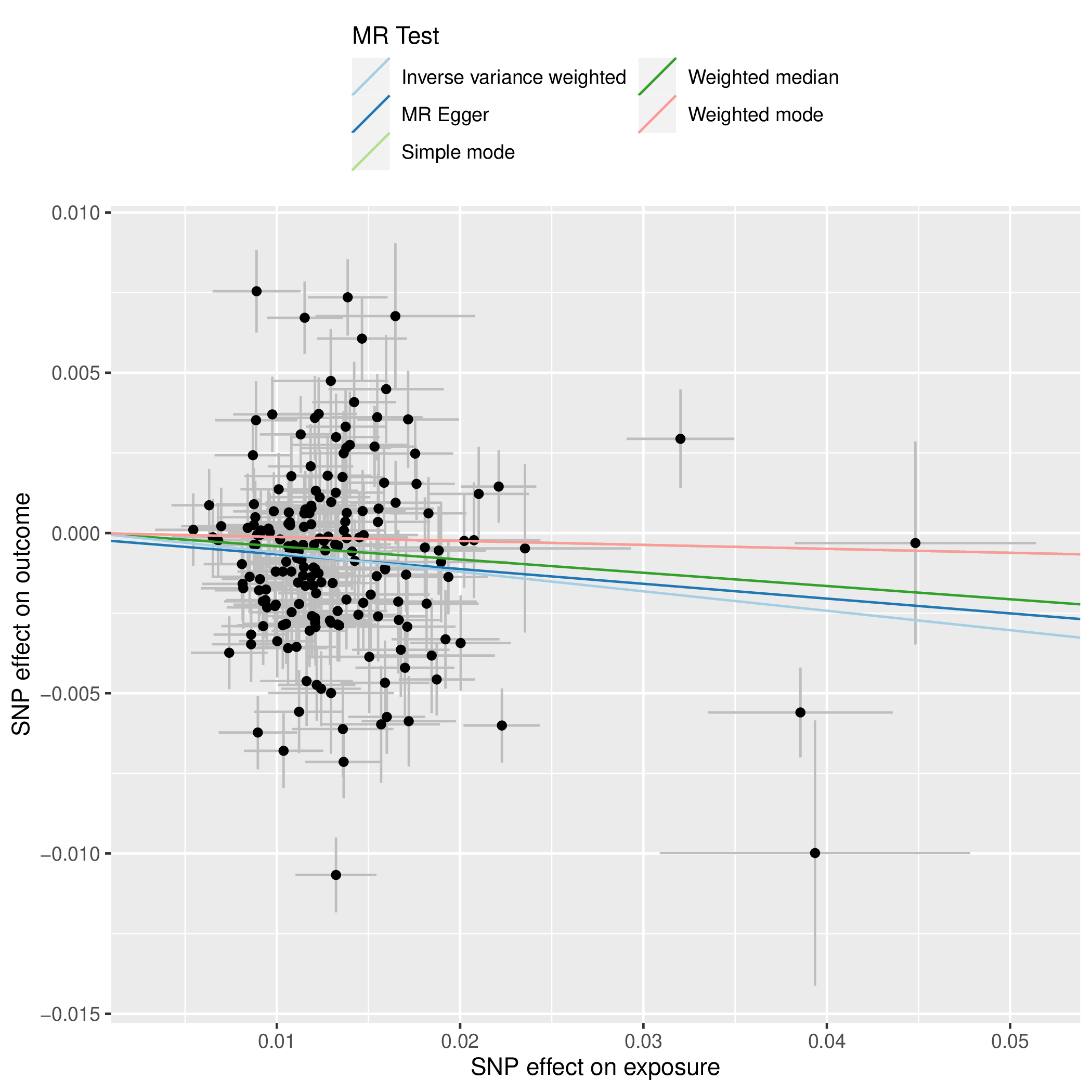

*Scatterplot showing the effect sizes of the SNP-exposure association and the SNP-outcome associations with standard error bars. The slopes of the lines correspond to causal estimates using each of the five different methods. All MR methods are in the same direction here.*

**Supplementary Figure S22.** Forest plot for smoking initiation as the exposure and executive function as the outcome.

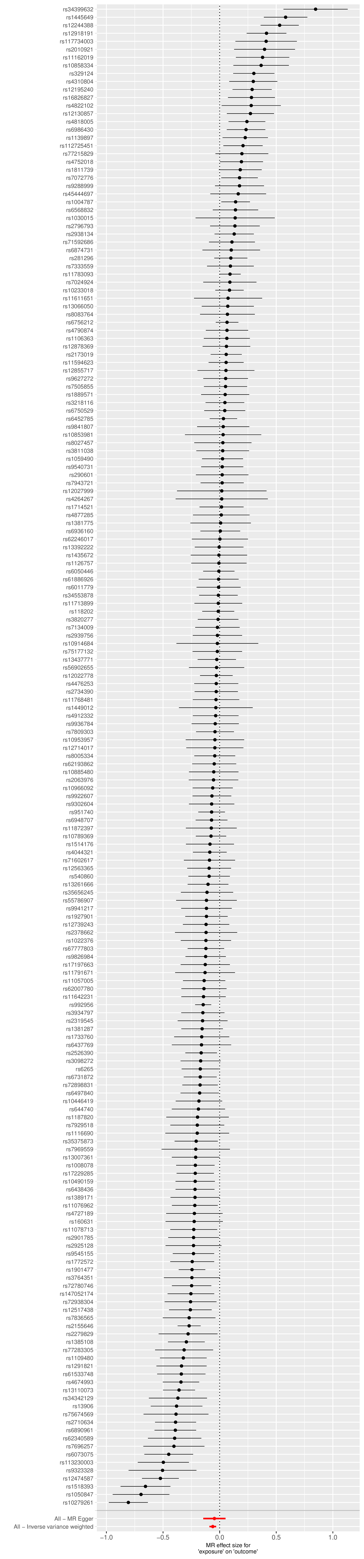

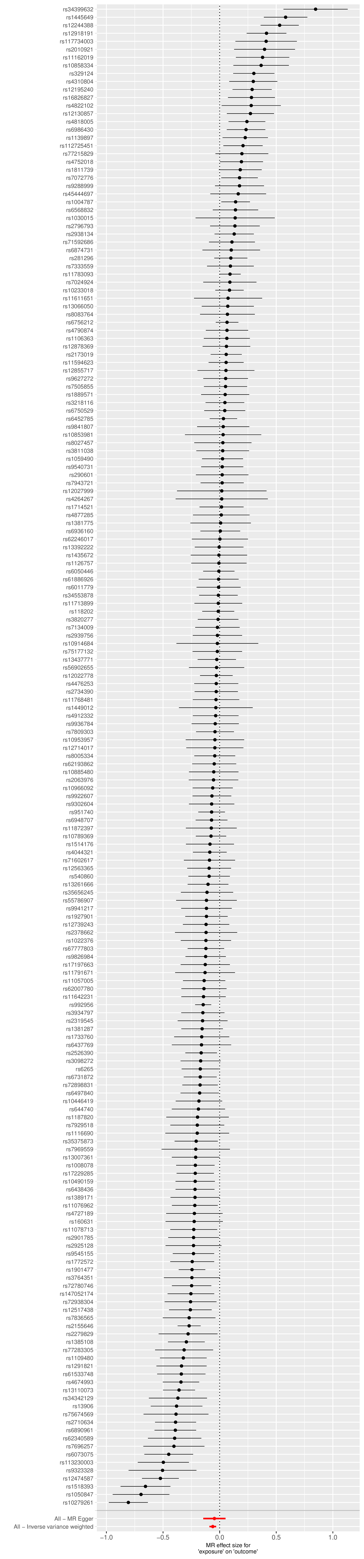

*Each black point represents the effect size for each SNP of the exposure on the outcome. Red points indicate the causal estimates for all SNPs as a single genetic instrument for the MR-Egger and the inverse-variance weighted (IVW) methods. Horizontal lines for each point represent 95% confidence intervals for each estimate. Some heterogeneity is observed here.*

**Supplementary Figure S23.** Funnel plot for smoking initiation as the exposure and executive function as the outcome.

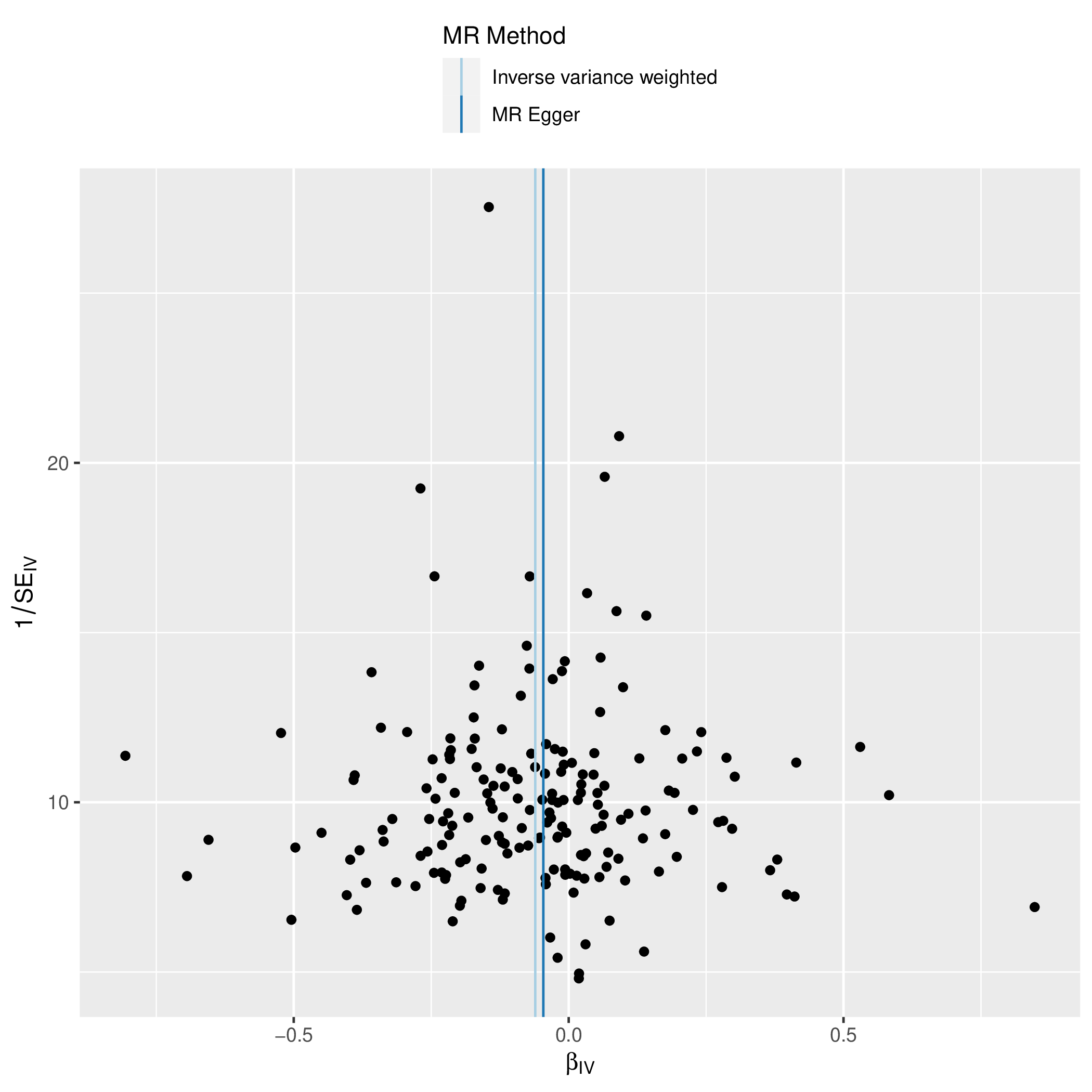

*Each SNP is used as a separate instrument to estimate a causal effect against the inverse of the standard error of that causal estimate. The vertical lines show the causal estimates for all SNPs using the IVW and MR-Egger methods. In this plot there is a degree of asymmetry, which may indicate violation of instrumental variable assumptions, e.g., horizontal pleiotropy.*

**Supplementary Figure S24.** Leave on out plot for smoking initiation as the exposure and executive function as the outcome.

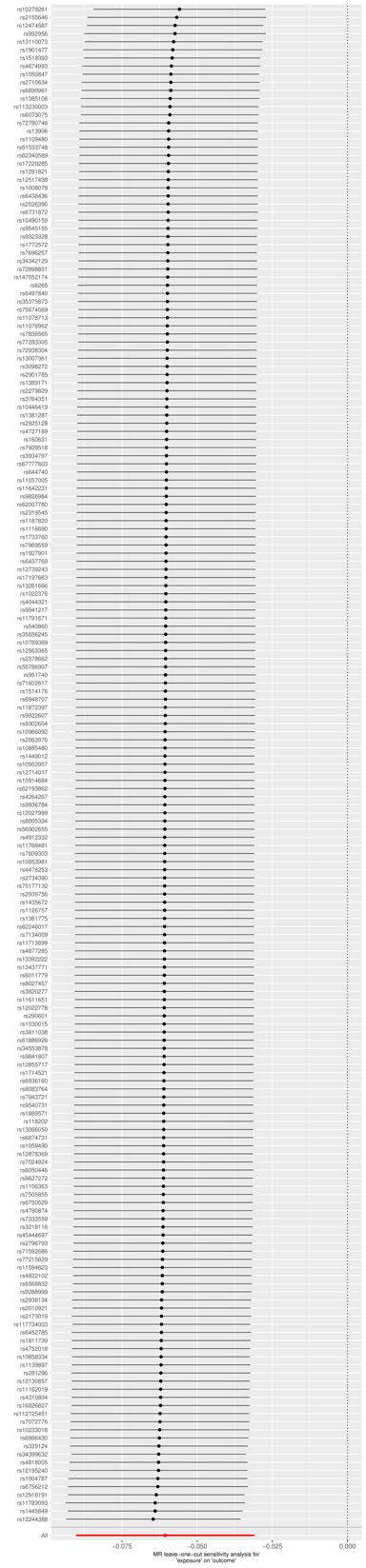

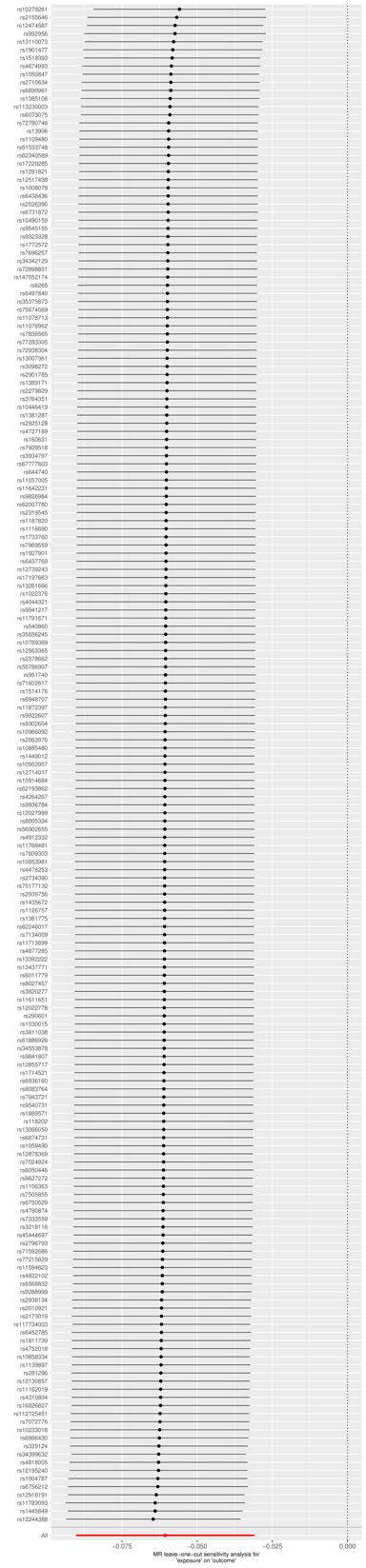

*Each black point represents the IVW estimate of the exposure on the outcome excluding that particular SNP from the analysis. The red point shows the IVW estimate using all SNPs in the instrument. There are no instances where the exclusion of one particular SNP leads to dramatic changes in the overall result.*
